# Cross-reactive neuraminidase inhibition antibodies against H5N1 by consecutive influenza A imprinting cohorts of the past century: population-based serosurvey, British Columbia, Canada

**DOI:** 10.1101/2025.09.19.25336209

**Authors:** Danuta M Skowronski, Charlene Ranadheera, Samantha E Kaweski, Suzana Sabaiduc, Lea Separovic, Gaby Makowski, Johnny Ung, Romina C Reyes, Bonnie Henry, Arianne Albert, Felicity Clemens, Darwyn Kobasa, Nathalie Bastien

## Abstract

**Background:** Avian influenza of the H5N1 subtype shares substantial relatedness in its neuraminidase (NA) surface protein with human influenza A H1N1 viruses of the past century. Understanding variation in pre-existing anti-N1 antibodies against H5N1 is critical to pandemic risk assessment and preparedness.

**Methods:** We used anonymized, residual sera collected equally from ten age groups spanning one to >80 years during an August 2024 cross-sectional serosurvey in British Columbia, Canada. We assessed NA inhibition antibody titres by enzyme-linked lectin assay against H5N1 (N=575), H1N1pdm09 (N=250) and H3N2 (N=205). We compared anti-NA titres by birth (imprinting) cohorts defined in relation to historic N1 and/or N2 exposure opportunities.

**Results:** Among participants with median age 32 (IQR: 15-62) years, 404 (70%) had cross-reactive anti-N1 titre >10 against H5N1, with 260 (45%), 182 (32%) and 98 (17%), having titres >40, >80 and >160, respectively. H5N1 titres were consistently lower but strongly associated with H1N1pdm09 (r=0.86; 95%CI:0.82-0.89). Geometric mean titres against H1N1pdm09 and H5N1 peaked among young adults born 1997-2003 (427.9, 100.8), declining to lows among young children born 2015-2023 (20.7, 6.8) and middle-aged adults born 1957-1967 (25.1, 10.7), increasing to similar secondary peak among older adults born pre-1947 (387.3, 81.0).

**Conclusions:** A substantial proportion of the population has pre-existing, cross-reactive anti-N1 antibodies against H5N1. We interpret variation by age and imprinting cohorts within a unifying hypothesis, emphasizing the role of historic influenza pandemics in expanding and refining the immune repertoire through heightened attack rates and shifts in immunological hierarchies. Our findings have implications for H5N1 and other zoonotic influenza risk assessment.

## Introduction

Over the past century, three hemagglutinin (HA) and two neuraminidase (NA) subtypes of influenza A have adapted to humans, causing the 1918 H1N1, 1957 H2N2, 1968 H3N2, 1977 H1N1 and 2009 H1N1pdm09 pandemics, each followed by seasonal epidemics as adapted viruses evolved to escape population immunity [1–4]. The H2N2 subtype displaced H1N1 and H3N2 displaced H2N2, but H1N1 and H3N2 viruses have co-circulated since 1977. Early periods in post-1918 H1N1 evolution were called “A-swine” (1918-33), “A0” (1934-46), and “A-prime” (1947-57) [3–6]. The 1977 H1N1 pandemic was caused by re-emergent “A-prime” virus and has been called a pseudo-pandemic because it mostly affected H1N1-inexperienced people <25 years [1,3]. The 2009 H1N1 pandemic was due to a complex H1N1pdm09 reassortment virus, more closely related to the “A-swine” era, and displaced “A-prime” ancestry strains [7]. Pre-school and school-aged children 1-4 and 5-19 years experienced peak 2009 pandemic attack rates (∼40% and 50%, respectively), declining successively thereafter among adults 20-44 (∼20%), 45-64 (∼15%), and >65 years (∼10%) [8].

The HA is more abundant than the NA on the influenza virus surface with the highly-exposed HA1 head typically immuno-dominant and subject to greater selection pressure and evolutionary mutation than the HA2 stalk or NA [1,9–11]. HA antibodies are virus neutralizing while NA antibodies are infection-permissive, restricting viral shedding and disease severity [1,11–13]. HA2 antibodies cross-react among subtypes belonging to the same phylogenetic group 1 (e.g., H1, H2, H5, H6) versus group 2 (e.g., H3, H4)[1,9]. Although NA subtypes are also separately categorized into group 1 (e.g., N1, N8) or group 2 (e.g., N2, N6), antibodies against the NA cross-react within the same subtype only [1,9,12]. In pairwise comparison of the percent identical amino acids, HA1 relatedness across historic human H1N1 viruses has ranged as low as 70-75%, higher within both the HA2 (>85%) and NA head (>80%) (**Table S1**) [1]. The H1N1 and H2N2 subtypes share even lower HA1 identity (<60%), with greater HA2 (>80%) than heterosubtypic NA (<50%) relatedness. Both group 1 subtypes share low HA1 (<40%) and HA2 (<60%) identities with H3N2 viruses, also low for heterosubtypic N1 (<50%), but high for homosubtypic N2 (>95% for H2N2 and H3N2 pandemic strains). These homo-group HA2 and/or homosubtypic NA relatedness and cross-reactivity considerations likely explain subtype replacement versus co-circulation following pandemics.

Through an immunological phenomenon called imprinting, the first influenza infection of childhood influences the lifelong immune response, attributed to the greater efficiency of memory responses to shared epitopes [1,14,15]. During major HA subtype shift (e.g., pandemics), the HA1 loses antigenic advantage, shifting immunologic hierarchies toward more conserved HA2 and/or NA epitopes [1,9,11]. NA responses are stronger when dissociated from the competitive HA, such as among immunologically naïve children or individuals imprinted to viruses with greater NA than HA identity [1,9,11,16]. Ultimately, however, even low-prevalence HA1-specific memory, such as accrued across seasonal exposures, reinstates HA1 immuno-dominance [14].

Recent expansion in the geographic and host range of highly pathogenic avian influenza of the H5N1 subtype, including newly affected mammals and associated human cases [1,17,18], has increased pandemic concern and renewed interest in potential population pre-immunity. Previous hypotheses have focused on potential cross-protective anti-HA2 effects [2], but a role for anti-N1 is reinforced by the high homosubtypic N1 identity (>85%) between H5N1 and human H1N1 strains, notably 1918 and 2009 pandemic viruses, like or exceeding HA2 identities (∼80-85%) (**Table S1**) [1]. To assess anti-N1 antibodies against H5N1 in the general population, we undertook a cross-sectional serosurvey, exploring variation by age and imprinting cohorts defined by N1 and/or N2 exposure opportunities over the past century.

## Methods

### Sample collection

We used a convenience sample of anonymized residual sera, collected within a ten-day period in August 2024 by LifeLabs, the only outpatient laboratory network in the most populated Lower Mainland region of British Columbia, Canada. Sera were provided under legal order of the Provincial Health Officer as part of COVID-19 seroprevalence monitoring [19], the latter sampling approach predicated on a longstanding influenza seroprevalence protocol [20–22]. Residual samples were selected and anonymized by the LifeLabs Central Processing Centre in consecutive order until 200 samples per age group, equally by sex, was reached (i.e., 0-4, 5-9, then 10-year bands through >80-years) (N=2000).

For testing of anti-N1 to H5N1, we initially randomly selected 50 specimens within each age group (n=500). We derived birth year as 2024 minus age at specimen collection. We defined 11 birth cohorts based upon subtype circulation, further considering age-related effects (e.g., 2009 pandemic incidences) (**Table 1**). To ensure at least 50 samples per birth cohort, we randomly selected additional sera as indicated, resulting in >50 for some age groups (N=575). For comparator H1N1pdm09 testing, we randomly selected 25 sera by birth cohort, combining the two youngest categories (10 cohorts, N=250), preferentially including sera with H5N1 results (n=229) and randomly selecting commensurate top-up as needed (n=21). Finally, we chose the four adult cohorts with lowest and highest H5N1 anti-N1 titres to also explore anti-N2 titres (N=205).

**Table 1.**
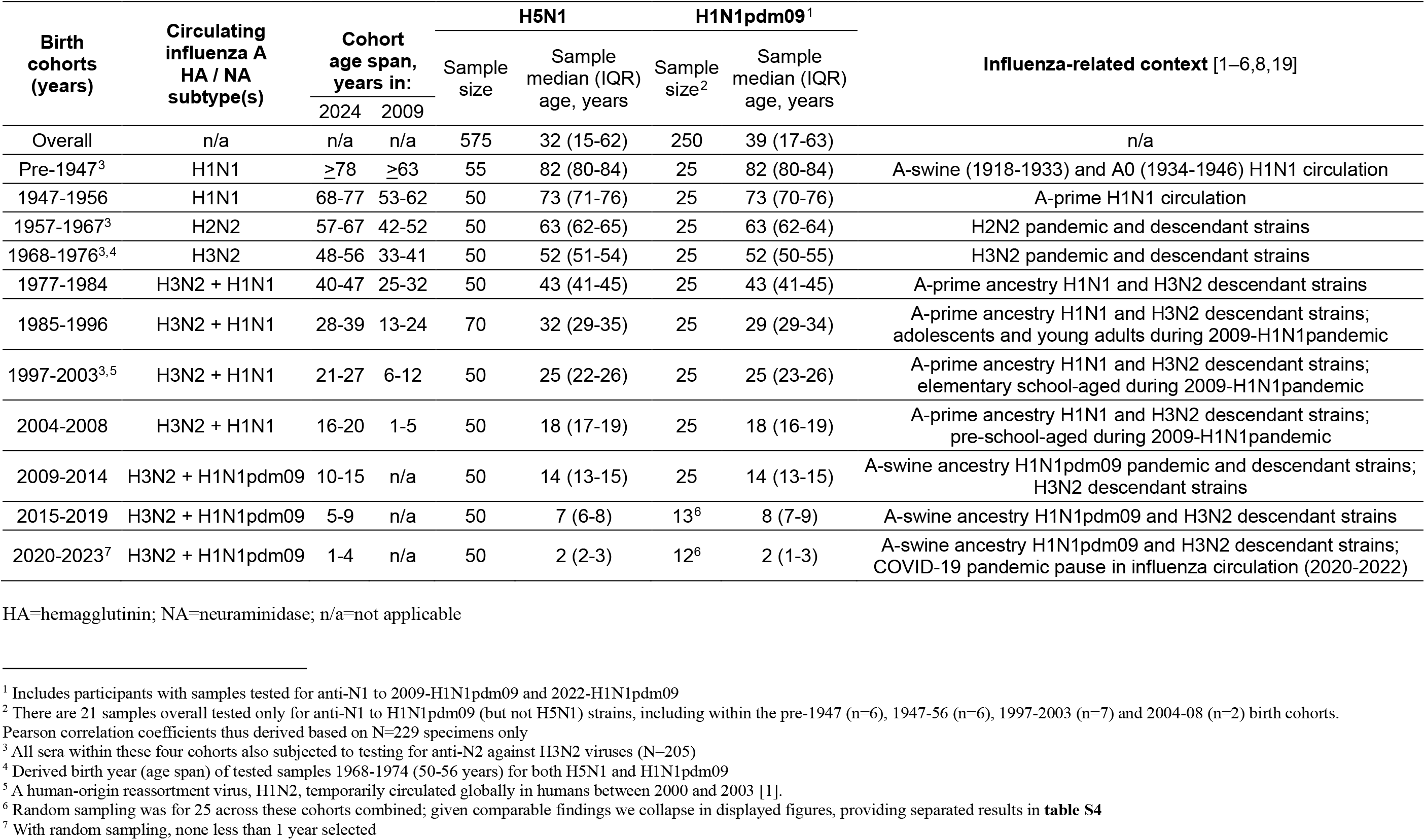
Sample size, median age and influenza-related context by birth cohort.

| Birth cohorts (years) | Circulating influenza A HA / NA subtype(s) | Cohort age span, years in: |  | H5N1 |  | H1N1pdm09<sup>1</sup> |  | Influenza-related context [1–6,8,19] |
| --- | --- | --- | --- | --- | --- | --- | --- | --- |
|  |  |  |  | Sample size | Sample median (IQR) age, years | Sample size<sup>2</sup> | Sample median (IQR) age, years |  |
|  |  | 2024 | 2009 |  |  |  |  |  |
| Overall | n/a | n/a | n/a | 575 | 32 (15-62) | 250 | 39 (17-63) | n/a |
| Pre-1947<sup>3</sup> | H1N1 | ≥78 | ≥63 | 55 | 82 (80-84) | 25 | 82 (80-84) | A-swine (1918-1933) and A0 (1934-1946) H1N1 circulation |
| 1947-1956 | H1N1 | 68-77 | 53-62 | 50 | 73 (71-76) | 25 | 73 (70-76) | A-prime H1N1 circulation |
| 1957-1967<sup>3</sup> | H2N2 | 57-67 | 42-52 | 50 | 63 (62-65) | 25 | 63 (62-64) | H2N2 pandemic and descendant strains |
| 1968-1976<sup>3,4</sup> | H3N2 | 48-56 | 33-41 | 50 | 52 (51-54) | 25 | 52 (50-55) | H3N2 pandemic and descendant strains |
| 1977-1984 | H3N2 + H1N1 | 40-47 | 25-32 | 50 | 43 (41-45) | 25 | 43 (41-45) | A-prime ancestry H1N1 and H3N2 descendant strains |
| 1985-1996 | H3N2 + H1N1 | 28-39 | 13-24 | 70 | 32 (29-35) | 25 | 29 (29-34) | A-prime ancestry H1N1 and H3N2 descendant strains; adolescents and young adults during 2009-H1N1pandemic |
| 1997-2003<sup>3,5</sup> | H3N2 + H1N1 | 21-27 | 6-12 | 50 | 25 (22-26) | 25 | 25 (23-26) | A-prime ancestry H1N1 and H3N2 descendant strains; elementary school-aged during 2009-H1N1pandemic |
| 2004-2008 | H3N2 + H1N1 | 16-20 | 1-5 | 50 | 18 (17-19) | 25 | 18 (16-19) | A-prime ancestry H1N1 and H3N2 descendant strains; pre-school-aged during 2009-H1N1pandemic |
| 2009-2014 | H3N2 + H1N1pdm09 | 10-15 | n/a | 50 | 14 (13-15) | 25 | 14 (13-15) | A-swine ancestry H1N1pdm09 pandemic and descendant strains; H3N2 descendant strains |
| 2015-2019 | H3N2 + H1N1pdm09 | 5-9 | n/a | 50 | 7 (6-8) | 13<sup>6</sup> | 8 (7-9) | A-swine ancestry H1N1pdm09 and H3N2 descendant strains |
| 2020-2023<sup>7</sup> | H3N2 + H1N1pdm09 | 1-4 | n/a | 50 | 2 (2-3) | 12<sup>6</sup> | 2 (1-3) | A-swine ancestry H1N1pdm09 and H3N2 descendant strains; COVID-19 pandemic pause in influenza circulation (2020-2022) |

HA=hemagglutinin; NA=neuraminidase; n/a=not applicable
<sup>1</sup> Includes participants with samples tested for anti-N1 to 2009-H1N1pdm09 and 2022-H1N1pdm09
<sup>2</sup> There are 21 samples overall tested only for anti-N1 to H1N1pdm09 (but not H5N1) strains, including within the pre-1947 (n=6), 1947-56 (n=6), 1997-2003 (n=7) and 2004-08 (n=2) birth cohorts. Pearson correlation coefficients thus derived based on N=229 specimens only
<sup>3</sup> All sera within these four cohorts also subjected to testing for anti-N2 against H3N2 viruses (N=205)
<sup>4</sup> Derived birth year (age span) of tested samples 1968-1974 (50-56 years) for both H5N1 and H1N1pdm09
<sup>5</sup> A human-origin reassortment virus, H1N2, temporarily circulated globally in humans between 2000 and 2003 [1].
<sup>6</sup> Random sampling was for 25 across these cohorts combined; given comparable findings we collapse in displayed figures, providing separated results in **table S4**
<sup>7</sup> With random sampling, none less than 1 year selected

### Test viruses

Anti-HA2 antibodies can sterically interfere with NA enzymatic activity, potentially over-estimating NA-specific antibody attribution [12,23]. Given lower cross-reactive anti-HA2 seroprevalence for group 2 than group 1 viruses, lowest against H7, we produced H7 reassortment viruses from A/Anhui/2013 (H7N9), the six internal proteins from A/PuertoRico/8/1934 (H1N1), and study virus Nas [24,25]. For H5N1, we used NA from a clade 2.3.4.4b H5N1 avian strain, A/RT-Hawk/ON/FAV-0473-4/2022. For H1N1pdm09, we used NA from a 2009 pandemic strain, A/InDRE/Mexico/4487/2009, and a 2022 descendant strain, A/Wisconsin/67/2022. For H3N2, we used a 1968 H3N2 pandemic strain, A/Hong Kong/1/1968, and a 2022 descendant strain, A/Massachusetts/18/2022. Test viruses were amplified in MDCK cells at Canada’s National Microbiology Laboratory under CL3; sequencing confirmed 100% pre-/post-amplification nucleotide identity. GISAID identifiers are provided in **Table S2**.

### Anti-NA assay

Functional neuraminidase inhibition (NAI) antibody testing was by enzyme-linked lectin assay (ELLA) under containment level 3 (CL3) as indicated [23,26]. Prior to testing, one part serum was treated with 4 parts 1/10 dilution of receptor destroying enzyme II (Accurate Chemical & Scientific Corporation), incubated 18h at 37°C and heat inactivated 8h at 56°C. Plates (96-well) were coated with 25µg/mL fetuin (stored up to 2 months at 4°C) and washed thrice with phosphate-buffered saline with 0.05% Tween-20 (PBS-T). Serially diluted sera were added to the fetuin-coated plates. Test viruses were diluted to 90% or 70% maximal ELLA signal for N1 or N2 viruses, respectively, with equal parts added to sera and incubated 20h at 37°C. Plates were washed 6x with PBS-T, and peanut agglutinin conjugated to horse-radish peroxidase (Millipore Sigma) was then added, incubating 2h at room temperature in darkness. Plates were washed thrice before adding o-phenylenediamine dihydrochloride (Millipore Sigma) and then incubated 10min at room temperature in darkness. An equal part 1N sulfuric acid was added to stop the reaction and plates were read at optical density 492nm with 650nm reference filter. Samples were run in duplicate and the mean of the two values used to determine the 50% inhibitory concentration (IC50) by variable slope (4-parameter) non-linear regression analysis (GraphPad Prism software). The inverse of the IC50 value is presented as the NAI titre, with titres below the lower limit of detection (i.e., <10) assigned a value of 5.

### Analyses

We calculated geometric mean titers (GMTs) with 95% confidence intervals (CIs) and compared across groups (e.g., age, sex, cohort, virus) using ANOVA (significance set at p<0.05). NAI correlates of protection have not been established but higher titres correspond with higher likelihood of protection [13,27,28]. We calculated group-specific percentages and 95%CIs with NAI titres >10 (detectable limit), >40, >80 and >160. We compared individual strain-specific titres through scatterplots, and calculated Pearson correlation coefficients with 95% CIs. Analyses used R Statistical Software (v4.2.3) on log2-transformed data.

### Ethics statement

The study was approved by University of British Columbia Clinical and Health Canada-Public Health Agency of Canada Research Ethics Boards.

## Results

### Anti-N1 seroprevalence

Main findings are displayed in **Figure 1** (GMTs) and **Figure 2** (percent meeting threshold titres), with counts and 95% CIs in **Table S3 (**age, sex) and **Table S4 (**birth cohorts). Among 575 participants with median age 32 (IQR: 15-62) years, 404 (70%) had detectable anti-N1 against H5N1, with titres >40, >80 and >160 in 260 (45%), 182 (32%) and 98 (17%), respectively. GMTs were consistently lower for H5N1 than H1N1pdm09; both varied by age and birth cohort (p<0.01), but not sex. The 2009-H1N1pdm09 and 2022-H1N1pdm09 titres were comparable for those born 2009-2014, higher against 2009-H1N1pdm09 among cohorts born pre-2009, but higher against 2022-H1N1pdm09 among cohorts born post-2015.

**Figure 1.**
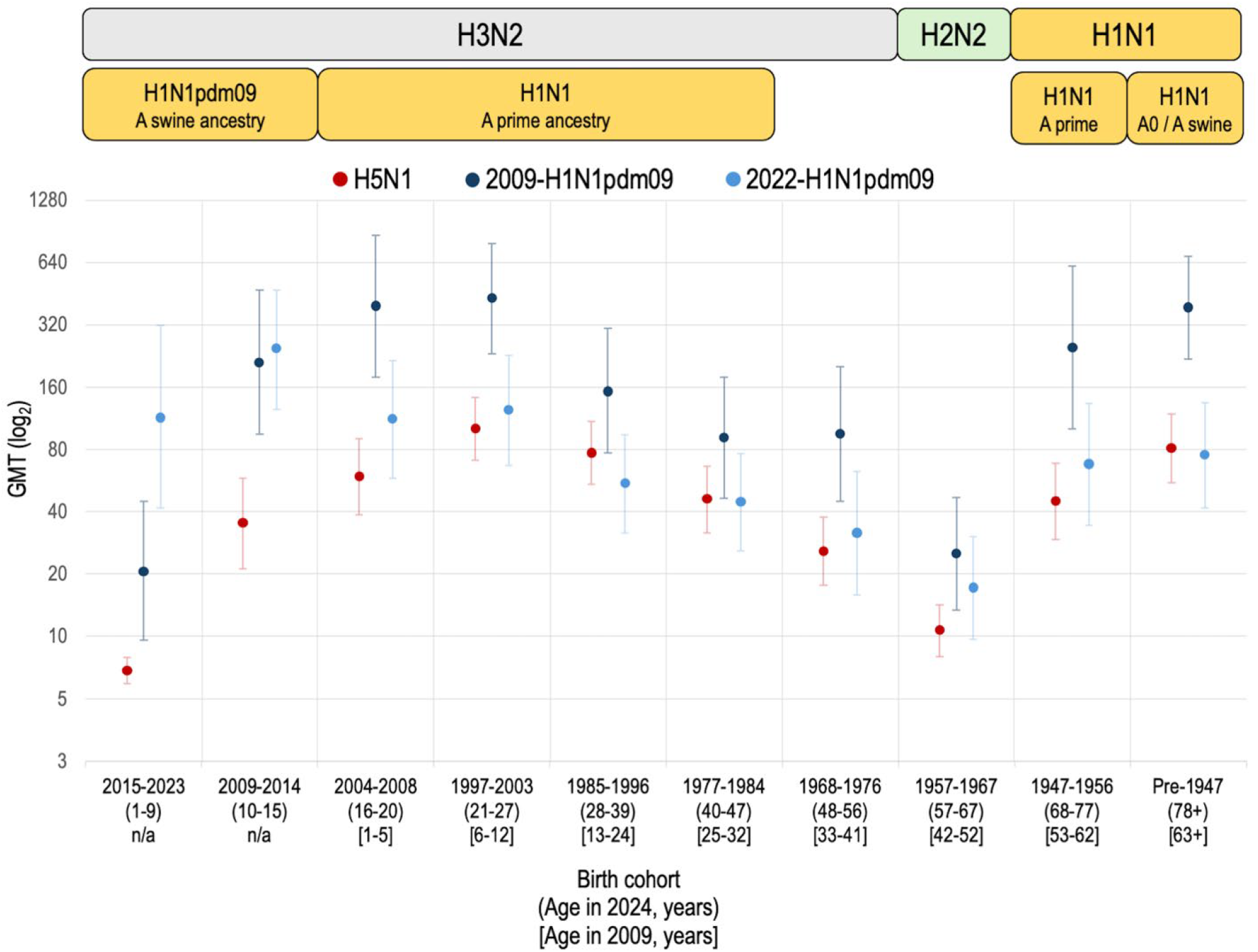
GMTs of NAI antibody against H5N1 and H1N1pdm09, by birth cohort GMT **=** geometric mean titre Displayed are GMTs and their 95% confidence intervals (CIs), by specified birth cohort (derived birth years), of neuraminidase inhibition (NAI) antibody against H5N1, 2009-H1N1pdm09 and 2022-H1N1pdm09 study viruses. All test viruses are H7 reassortment strains. Corresponding epochs of influenza A subtype circulation are annotated above the figure. Sample sizes by cohort and influenza A test virus are provided in **Table 1**. Precise values with 95% confidence intervals are provided in **Table S4**. Anti-N1 GMTs against H5N1 were comparably low (<10) for the youngest 2020-2023 and next youngest 2015-19 birth cohorts shown collapsed here but also stratified in **Table S4**.

**Figure 2.**
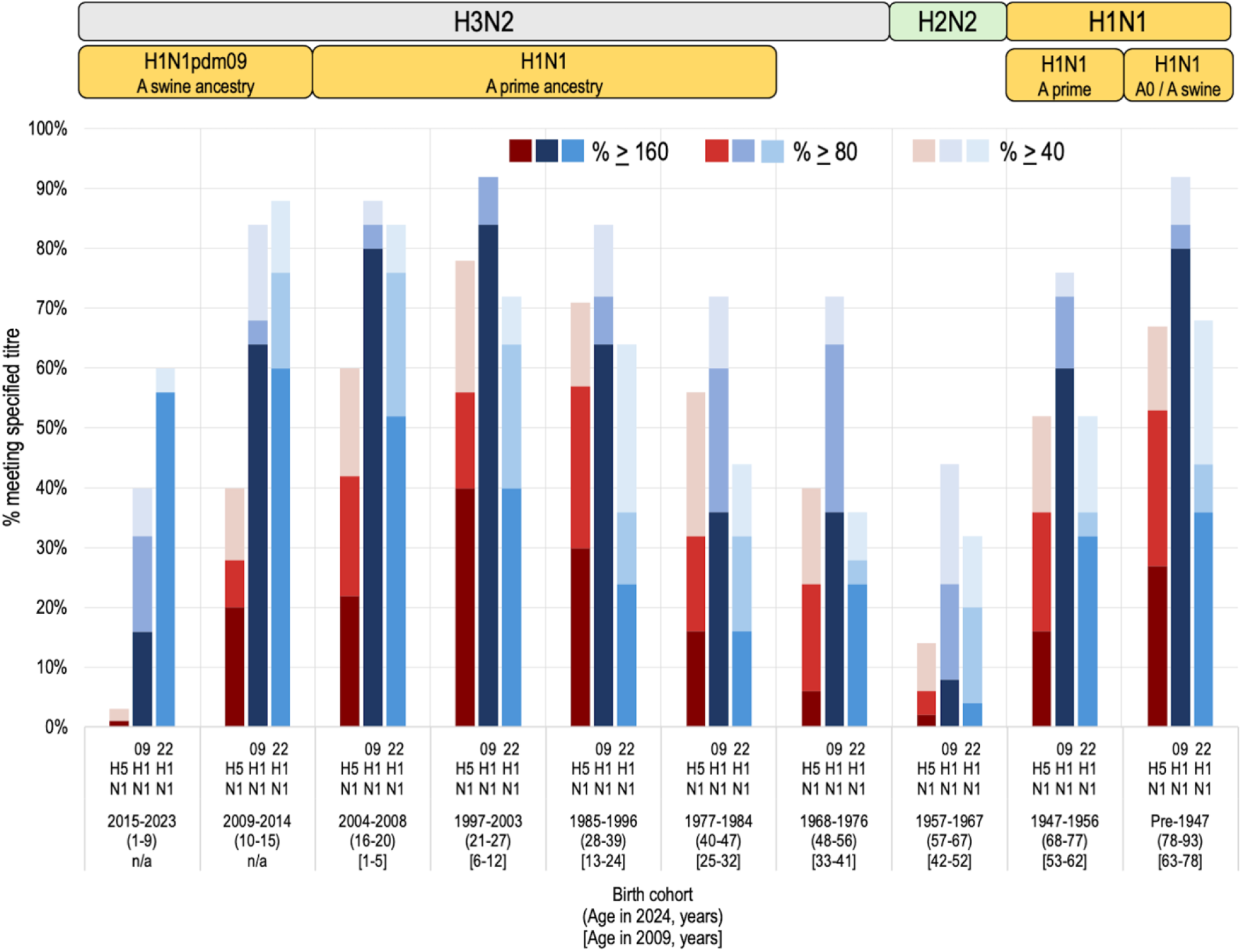
Percent meeting NAI threshold titres against H5N1 and H1N1pdm09, by birth cohort Displayed are the percentage meeting specified threshold titres of neuraminidase inhibition (NAI) antibody against H5N1 in shades of red, and 2009-H1N1pdm09 (‘09 H1N1) or 2022-H1N1pdm09 (‘22 H1N1) in shades of blue. All test viruses are H7 reassortment strains. Corresponding epochs of influenza A subtype circulation are annotated above the figure. Sample sizes by cohort and influenza A test virus are provided in **Table 1**. Precise values with 95% confidence intervals are provided in **Table S4**. Anti-N1 proportions for H5N1 were comparably low (<6%) for the youngest 2020-2023 and next youngest 2015-2019 birth cohorts, shown collapsed here but also stratified in **Table S4**.

Three birth cohorts had the highest anti-N1 GMTs to 2009-H1N1pdm09 and amongst the highest against H5N1, without significant difference between them for either virus, including (youngest to oldest): 2004-2008 (394.8 and 59.1); 1997-2003 (427.9 and 100.8); and pre-1947 (387.3 and 81.0). At least 80% within these respective cohorts had titres >160 to 2009-H1N1pdm09 (80%, 84%, 80%) and most had titre >40 to H5N1 (60%, 78%, 67%) including a substantial proportion with H5N1 titre >160 (22%, 40%, 27%). Anti-N1 titres within the younger adjacent 2009-2014 cohort did not differ significantly from the above three cohorts for 2009-H1N1pdm09 (211.5, 64% >160) but were significantly lower than the 1997-2003 (p<0.001) and pre-1947(p=0.01) cohorts for cross-reactive H5N1 (35.1, 40% >40). Conversely, the older adjacent 1985-1996 cohort had significantly lower 2009-H1N1pdm09 titres (153.6, 64% >160) than the 1997-2003 (p=0.027) and pre-1947 (p=0.039) cohorts, but did not differ significantly from either cohort for H5N1 (77.0, 71% >40).

Three birth cohorts had anti-N1 GMTs against 2009-H1N1pdm09 and H5N1 that were significantly lower than each of the above three highest cohorts (p<0.01), including (youngest to oldest): 2015-2023 (20.7 and 6.8), 1968-1976 (95.1 and 25.7), and 1957-1967 (25.1 and 10.7). For 2009-H1N1pdm09, the 2015-2023 and 1957-1967 cohorts did not differ significantly from each other, but both were significantly lower than the 1968-1976 cohort (p<0.01); for H5N1 all three pairwise comparisons were significantly different (p<0.01). For 2009-H1N1pdm09, a minority within each respective cohort had anti-N1 titre >160 (16%, 36%, 8%). For H5N1, a minority had titre >40 (3%, 40%, 14%) including a negligible proportion with H5N1 titre >160 (1%, 6%, 2%). Relative to the 1968-1976 cohort, the adjacent 1977-1984 cohort had similar 2009-H1N1pdm09 (90.9) but significantly higher H5N1 GMT (45.7) (p=0.033); the 1977-1984 GMTs were lower than the 1947-1956 cohort (both H1N1 A-prime ancestry epochs) for 2009-H1N1pdm09 (248.2) but similar for H5N1 (44.7), with neither difference significant.

Scatterplots reinforce that anti-N1 titres against H5N1 are strongly correlated with anti-N1 titres against 2009-H1N1pdm09, with correlation coefficient of 0.86 (95%CI: 0.82-0.89) overall and >0.70 by birth cohort except 2015-2023 (0.64; 95%CI: 0.32-0.82) (**Figure 3**). Correlation coefficients were significantly lower overall between H5N1 and 2022-H1N1pdm09 at 0.52 (95%CI: 0.42-0.61) overall, also lower by cohort including 2015-2023 (0.50; 95%CI: 0.13-0.75), noting wider confidence intervals with cohort stratification (**Figure S1**).

**Figure 3.**
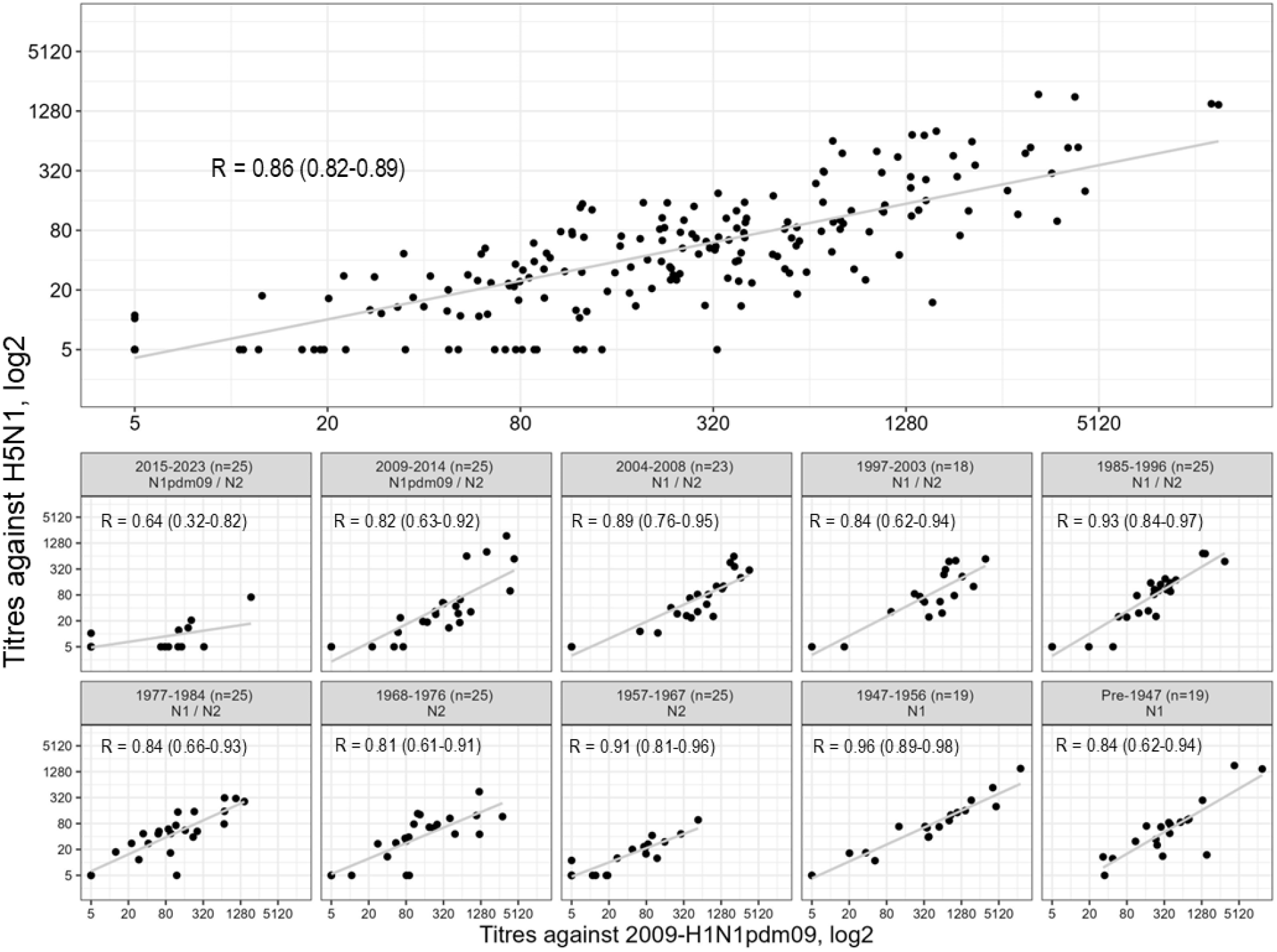
Scatterplot of relationship between NAI titres against H5N1 and 2009-H1N1pdm09 Scatterplots display the relationship between H5N1 and 2009-H1N1pdm09 neuraminidase inhibition (NAI) antibody titres, overall and by birth cohort, with accompanying Pearson correlation coefficients (r). Both test viruses are H7 reassortment strains. Within panel headers by birth cohort are displayed the range of cohort birth years (sample size) and circulating neuraminidase subtype(s) during the specified period. Only individuals tested for both viruses are included (n=229): the 21 samples tested only for anti-N1 to H1N1pdm09 but not H5N1 strains are excluded from the pre-1947 (n=6), 1947-56 (n=6), 1997-2003 (n=7) and 2004-08 (n=2) birth cohort scatterplots.

### Anti-N2 seroprevalence

Anti-N2 findings within the two adult cohorts with lowest (1957-1967, 1968-1976) and highest (1997-2003, pre-1947) anti-N1 GMTs against H5N1 are shown in **Figure 4**, with precise values provided in **Tables S5-S6**. Within these four adult cohorts, 1968-H3N2 exceeded 2022-H3N2 titres, and we focus on the former. The 1957-1967 cohort with lowest anti-N1 had highest anti-N2 GMT (642.7), significantly exceeding 2009-H1N1pdm09 (>25-fold) and H5N1 (>60-fold) GMTs (p<0.001). Conversely, the 1997-2003 cohort with highest anti-N1 had lowest anti-N2 GMT (31.5), significantly lower than 2009-H1N1pdm09 (>13-fold) and H5N1 (>3-fold) (p<0.001). While the preceding might suggest inverse N1:N2 relationship, the pre-1947 cohort with next-highest anti-N1, also had next-highest anti-N2 GMT (279), comparable to 2009-H1N1pdm09 (ratio=1.4) but significantly greater than H5N1 (3.5-fold) (p<0.001). In fact, the proportion with anti-N2 >160 was high for both the 1957-1967 and pre-1947 cohorts (100% and 78%), and all within the pre-1947 cohort also had anti-N2 >40. Finally, the 1968-1976 cohort with next-lowest anti-N1 also had next-lowest anti-N2 GMT (89.0), comparable to 2009-H1N1pdm09 (ratio=1.1) but also significantly greater than H5N1 (3.5-fold) (p<0.01). Overall, there was no N1:N2 relationship, with correlation coefficients comparing 1968-H3N2 to 2009-H1N1pdm09 of -0.06 (95%CI: -0.27-0.15) and to H5N1 of -0.14 (95%CI: -0.27-0.50) (**Figures S2-S3**).

**Figure 4.**
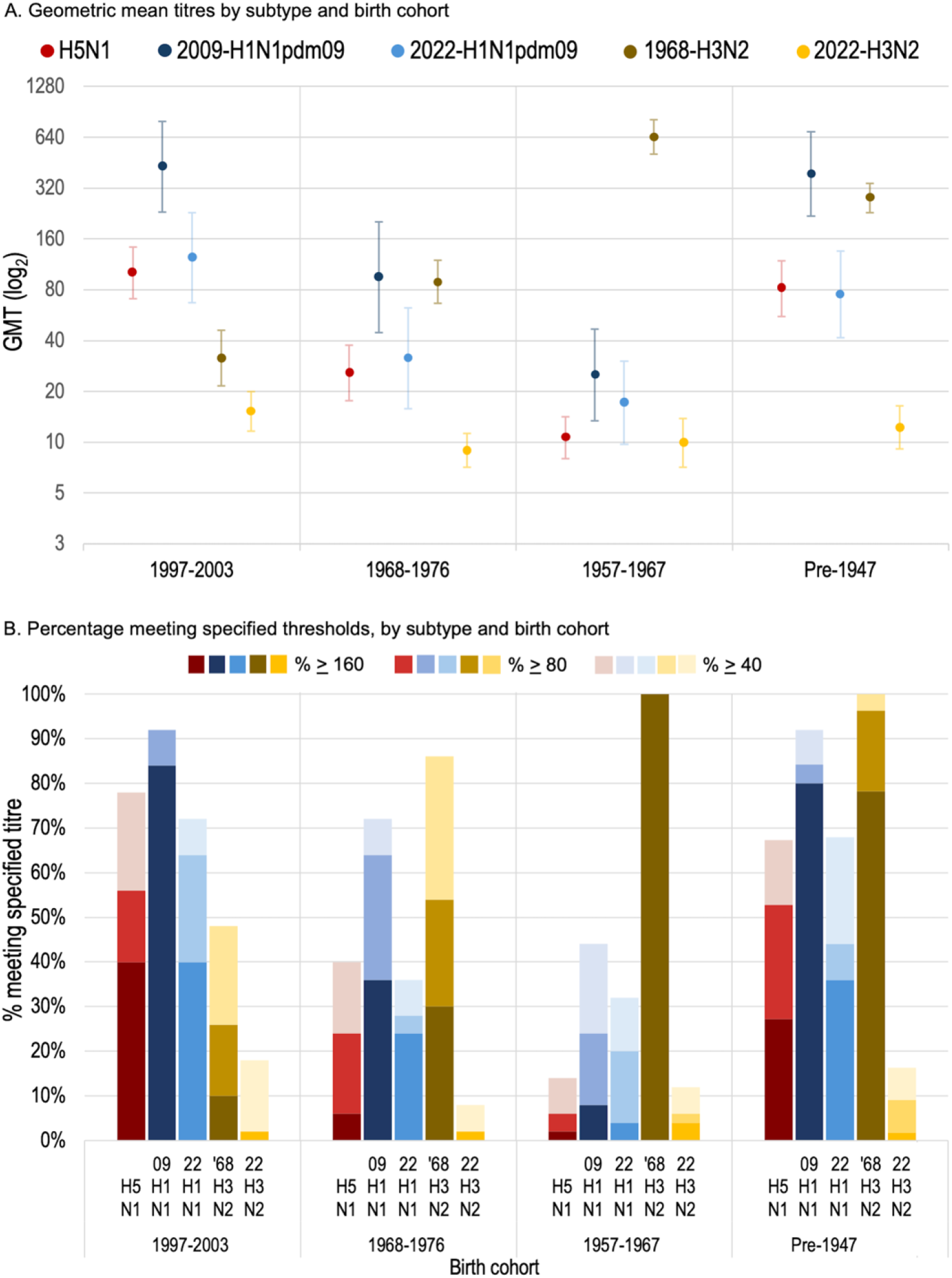
GMTs and percent meeting NAI threshold titres against H5N1, H1N1pdm09 and H3N2, by birth cohort For the four adult birth cohorts with two lowest (1957-1967, 1968-1976) and two highest (1997-2003, pre-1947) neuraminidase inhibition (NAI) antibody titres against H5N1, we display: (A) geometric mean titres (GMTs) and their 95% confidence intervals (CIs) and (B) percentage meeting specified thresholds titres including against H5N1, 2009-H1N1pdm09, 2022-H1N1pdm09, 1968-H3N2, and 2022-H3N2 study viruses. All test viruses are H7 reassortment strains. Samples tested for ant-N1 against H5N1 and anti-N2 against H3N2 include 50 per birth cohort except within the oldest (pre-1947: n=55) cohort; samples tested against each of the H1N1pdm09 viruses include 25 per birth cohort. For the 1997-2003 and pre-1947 cohorts, H1N1pdm09 findings were similar with restriction to the 18/25 and 19/25 samples, respectively, also tested against H5N1 and H3N2. Precise values are provided in **Tables S5-S6**.

## Discussion

With H5N1 expansion but without endemicity established in humans anywhere globally to date, we report cross-reactive anti-N1 antibodies to this emerging zoonosis in more than two-thirds of participants aged one to >80 years, including half with titre >40, one-third >80 and nearly 20% >160. Cross-reactive H5N1 and 2009-H1N1pdm09 titres were strongly correlated, peaking among young adults born 1997-2003, declining to lows among the youngest children born 2015-2023 and middle-aged adults born 1957-1967, thereafter increasing to a similar secondary peak among the oldest adults born pre-1947. We interpret variation by age and imprinting cohorts within a unifying hypothesis, emphasizing the role of historic influenza pandemics in expanding and refining the immune repertoire through heightened attack rates and shifts in immunological hierarchies.

Peak anti-N1 titres within the 1997-2003 birth cohort is consistent with having been school-aged children (6-12-years-old) during the 2009 pandemic, collectively experiencing the highest H1N1pdm09 attack rates [8]. Using sera from 130 participants aged 5-70 years, Daulagala et al explored age-dependent variation in NAI titres against historic H1N1 strains, also showing peak among cohorts aged 6-12-years during the 2009 pandemic [29]. Similar age-related considerations apply to the adjacent 2004-2008 and 1985-1996 cohorts with next-highest 2009 pandemic incidences [8]. Lower titres among successively younger post-2009 pediatric cohorts align with lower seasonal attack rates and exposure opportunities. With mounting HA1 memory, seasonal epidemics are also progressively less likely to back-boost existing anti-NA, also potentially more narrowly focused. These combined effects may be exemplified within the youngest 2015-2023 cohort with higher 2022-than 2009-H1N1pdm09 titres and negligible H5N1 cross-reactivity. On opposite side of the peak, successively lower anti-N1 among successively older cohorts reflects successively lower 2009 pandemic incidences with age, notwithstanding accumulated seasonal exposures to A-prime ancestry strains. Such age-related hypothesis alone, however, doesn’t explain the anti-N1 trough we observe within the 1957-1967 H2N2-imprinted cohort, significantly lower than both the younger 1968-1976 H3N2-imprinted and older pre-1957 H1N1-imprinted cohorts.

To further refine understanding through an imprinting lens, we incorporate intra-virionic antigenic competition, differential HA2 and NA relatedness, and shifts in immunological hierarchies associated with major HA1 change during pandemics [1,16,30,31]. The 1957-1967 H2N2-imprinted cohort had low anti-N1 but high anti-N2 titres, suggesting inverse N1:N2 relationship, but we also observed high anti-N2 in the pre-1947 cohort with high anti-N1 titres. A smaller 2020 study also showed substantial anti-N2 titres among people born during the pre-1957 H1N1 era (n=45), comparable to 1957-1968 (n=13) [32]. Our scatterplots showed no heterosubtypic NA correlation, reinforcing independence of subtype-specific NA cross-reactivity. With far lower heterosubtypic NA identity (<50%), the H2N2 subtype shares substantial homo-group HA2 identity (>80%) with both H1N1 and H5N1. Upon H1N1 exposure (or re-exposure), we postulate the H2N2-imprinted cohort retains hierarchical HA2 predilection, preferentially recalling memory responses to more conserved HA2 over de novo or even back-boosted NA response. This context differs from older cohorts lacking prior H2N2 infection since H1N1 viruses of the past century share more comparable HA2 and NA identities, reducing intra-virionic competition. Our hypothesis also explains the very high anti-N2 titres within the 1957-1967 cohort: with shift from group 1 H2 to group 2 H3 during the 1968 pandemic the opposite configuration of low HA2 but high N2 identity (<60% versus >95%) supports prominent anti-N2 back-boost, while also being permissive of anti-N2 responses in older cohorts.

Our findings align with other experimental and epidemiological evidence. Anti-N1 antibodies, notably those induced by H1N1pdm09, cross-react against H5N1 and protect animals from H5N1 challenge [11,13,33–36]. Our recent pre-print re-analysis of human zoonotic case reports shows lower H5N1 case fatality ratios post-2009 and, despite highly conserved H5 stalk identity, fewer adults >65 years among H5N1 versus H5N6 cases, consistent with anti-N1 as added protective factor [1]. Other smaller studies report anti-N1 seroprevalence against H5N1. This includes two publications involving blood donors with sample collection from Thailand in 2013 (n=100) [37] or Hong Kong in 2000 (n=50) and 2020 (n=63) [38], and a third publication involving United States (US) military personnel in 2000 (n=100) and 2008 (n=100) [39]. None of these publications included children. Two recent preprints reporting US serological studies include children among US Flu VE network participants in 2023-2024 (n=234, with 19 children 0.5-3 years) [40], and another using variously sourced samples from 2005 and 2017 (n=243, up to 36 children 1-17 years) [41]. Most participants in the above studies had detectable anti-N1 against H5N1, also correlated with H1N1pdm09, with potential imprinting effects raised in recent preprints. None of the studies, however, was able to directly compare anti-N1 titres between consecutive birth cohorts fully spanning the past century or to interpret variation within a unifying hypothesis incorporating both age and imprinting effects. In attempting to mitigate anti-HA2 effects on anti-NA findings, we also note these studies used group 1 H1 [39] or H6 [38,40,41], or group 2 H4 [37] reassortment test viruses, each with higher cross-reactive seroprevalence than the group 2 H7 reassortment we used [25]. The extent to which residual anti-HA2 effects may influence inter-cohort or cross-study comparisons is unclear.

The greatest limitation of anti-NA serosurveys is the lack of threshold correlates of protection. We cannot infer immunity based upon anti-NA levels, but patterns indicate higher likelihood of N1-based protection among young adults, aligned with observations of mild H5N1 illness in US animal workers [42]; and among pre-1947 birth cohorts, aligned with the paucity of older adult H5N1 cases globally [1]. As elsewhere, our anti-N1 findings for H5N1 represent cross-reactive over case-specific detection and we did not assess cross-neutralizing antibodies that are mostly HA-driven and identified at only low frequency even among animal workers [40,43]. We used a single clade 2.3.4.4b H5N1 virus but the NA of H5N1 viruses is highly conserved (**Table S1**) [1], with similar anti-N1 findings reported by others regardless of clade or genotype [38–41]. Influenza immunity is complex and other components (e.g., anti-HA2, T-cell) require consideration; systematic comparison of anti-HA2 titres between 1957-1967 and flanking cohorts could be of particular interest. Our anonymized approach precludes individual linkage to infection or vaccine history, but in addition to infections, older cohorts may have accrued more seasonal influenza vaccination. Although influenza infection clearly induces anti-NA responses, the same cannot be asserted for vaccination: the NA content of vaccines is not standardized and among influenza-experienced individuals, current seasonal formulations are mostly expected to induce HA response [28,33,40,44,45]. Our birth cohort definitions did not incorporate potential 10-year delay to first childhood influenza infection [46]: some within the 1947-1956 cohort may have acquired first infection post-1957, and some born 1957-1967 may not have been infected until post-1968; absent such misclassification, the 1957-1967 trough we report could be more pronounced. Finally, additional large seroprevalence studies are needed to confirm the robustness and generalizability of our findings.

In conclusion, we show a substantial proportion of the general population with pre-existing anti-N1 against zoonotic H5N1, potentially mitigating the pandemic risk despite expansion among other, presumably immunologically naïve, mammals. People with highest probability of anti-N1 cross-protection include young adults born during the 1990s or early 2000s likely due to their high incidence of H1N1pdm09 infection as schoolchildren during the 2009 pandemic, and older adults born pre-1947 likely due to closely related ancestral H1N1 imprinting and accumulated back-boost opportunities. Conversely, those with lowest likelihood of anti-N1 cross-protection include children born post-2009 with least influenza experience and middle-aged adults born during the 1957-1967 H2N2-imprinting epoch. Whether anti-N1 findings explain H5N1 mildness among animal workers or scarcity among older adults or suggest prioritized protective measures for young children and middle-aged adults, warrants further investigation. Our unifying hypothesis, incorporating both age (incidence) and imprinting (immunological hierarchy) effects of pandemics has implications for H5N1 and other zoonotic influenza risk assessment.

## Data Availability

Aggregate serological data are provided within the manuscript and supplementary material. Any further data sharing of seroprevalence data will be considered upon reasonable request to the corresponding author with appropriate review and aggregation, as required to comply with the provincial legislation under which the data were assembled and respecting privacy and confidentiality requirements.

## Author contributions

Conception and design: DMS, CR, SEK, SS, NB; acquisition of data: SEK, GM, JU; analysis of data: DMS, SEK, LS, AA, FC; laboratory supervision: CR, RCR, DK, NB; interpretation of data: DMS, CR, SEK, SS, BH, NB; drafting manuscript: DMS, SEK; critical review of manuscript: all; approval to publish: all.

## References

1. Skowronski DM, Kaweski SE, Separovic L, et al. Neuraminidase imprinting and the agerelated risk of zoonotic influenza. medRxiv preprint 2025; Available at: http://medrxiv.org/lookup/doi/10.1101/2025.07.03.25330844. Accessed 6 July 2025.

2. Gostic KM, Ambrose M, Worobey M, Lloyd-Smith JO. Potent protection against H5N1 and H7N9 influenza via childhood hemagglutinin imprinting. Science 2016; 354:722–726.

3. Kilbourne ED. Influenza pandemics of the 20th century. Emerg Infect Dis 2006; 12:9–14.

4. Zimmer SM, Burke DS. Historical Perspective — Emergence of Influenza A (H1N1) Viruses. N Engl J Med 2009; 361:279–285.

5. Budd AP, Beacham L, Smith CB, et al. Birth Cohort Effects in Influenza Surveillance Data: Evidence That First Influenza Infection Affects Later Influenza-Associated Illness. J Infect Dis 2019; 220:820–829.

6. Monto AS, Malosh RE, Petrie JG, Martin ET. The Doctrine of Original Antigenic Sin: Separating Good From Evil. J Infect Dis 2017; 215:1782–1788.

7. Morens DM, Taubenberger JK, Fauci AS. The persistent legacy of the 1918 influenza virus. N Engl J Med 2009; 361:225–229.

8. Van Kerkhove MD, Hirve S, Koukounari A, Mounts AW, H1N1pdm serology working group. Estimating age-specific cumulative incidence for the 2009 influenza pandemic: a metaanalysis of A(H1N1)pdm09 serological studies from 19 countries. Influenza Other Respir Viruses 2013; 7:872–886.

9. Kosik I, Yewdell JW. Influenza Hemagglutinin and Neuraminidase: Yin–Yang Proteins Coevolving to Thwart Immunity. Viruses 2019;

10. Kirkpatrick E, Qiu X, Wilson PC, Bahl J, Krammer F. The influenza virus hemagglutinin head evolves faster than the stalk domain. Sci Rep 2018; 8:10432.

11. Eichelberger MC, Wan H. Influenza Neuraminidase as a Vaccine Antigen. In: Oldstone MBA, Compans RW, eds. Influenza Pathogenesis and Control - Volume II. Cham: Springer International Publishing, 2015: 275–299. Available at: 10.1007/82_2014_398.

12. Krammer F, Fouchier RAM, Eichelberger MC, et al. NAction! How Can Neuraminidase-Based Immunity Contribute to Better Influenza Virus Vaccines? mBio 2018; 9:e02332–17.

13. Memoli MJ, Shaw PA, Han A, et al. Evaluation of Antihemagglutinin and Antineuraminidase Antibodies as Correlates of Protection in an Influenza A/H1N1 Virus Healthy Human Challenge Model. mBio 2016; 7:e00417–00416.

14. Knight M, Changrob S, Li L, Wilson PC. Imprinting, immunodominance, and other impediments to generating broad influenza immunity. Immunol Rev 2020; 296:191–204.

15. Gostic KM, Bridge R, Brady S, Viboud C, Worobey M, Lloyd-Smith JO. Childhood immune imprinting to influenza A shapes birth year-specific risk during seasonal H1N1 and H3N2 epidemics. PLoS Pathog 2019; 15:e1008109.

16. Kilbourne ED, Cerini CP, Khan MW, Mitchell JW, Ogra PL. Immunologic response to the influenza virus neuraminidase is influenced by prior experience with the associated viral hemagglutinin. I. Studies in human vaccinees. J Immunol 1987; 138:3010–3013.

17. Krammer F, Hermann E, Rasmussen AL. Highly pathogenic avian influenza H5N1: history, current situation, and outlook. Journal of Virology 2025; 99:e02209–24.

18. Peacock T, Moncla L, Dudas G, et al. The global H5N1 influenza panzootic in mammals. Nature 2024;

19. Skowronski DM, Kaweski SE, Irvine MA, et al. Serial cross-sectional estimation of vaccine-and infection-induced SARS-CoV-2 seroprevalence in British Columbia, Canada. CMAJ 2022; 194:E1599–E1609.

20. Skowronski DM, Hottes TS, McElhaney JE, et al. Immuno-epidemiologic correlates of pandemic H1N1 surveillance observations: higher antibody and lower cell-mediated immune responses with advanced age. J Infect Dis 2011; 203:158–167.

21. Skowronski DM, Hottes TS, Janjua NZ, et al. Prevalence of seroprotection against the pandemic (H1N1) virus after the 2009 pandemic. CMAJ 2010; 182:1851–1856.

22. Skowronski DM, Moser FS, Janjua NZ, et al. H3N2v and other influenza epidemic risk based on age-specific estimates of sero-protection and contact network interactions. PLoS One 2013; 8:e54015.

23. Couzens L, Gao J, Westgeest K, et al. An optimized enzyme-linked lectin assay to measure influenza A virus neuraminidase inhibition antibody titers in human sera. Journal of Virological Methods 2014; 210:7–14.

24. Neumann G, Watanabe T, Ito H, et al. Generation of influenza A viruses entirely from cloned cDNAs. Proceedings of the National Academy of Sciences 1999; 96:9345–9350.

25. Nachbagauer R, Choi A, Hirsh A, et al. Defining the antibody cross-reactome directed against the influenza virus surface glycoproteins. Nat Immunol 2017; 18:464–473.

26. Lambré CR, Terzidis H, Greffard A, Webster RG. Measurement of anti-influenza neuraminidase antibody using a peroxidase-linked lectin and microtitre plates coated with natural substrates. Journal of Immunological Methods 1990; 135:49–57.

27. Couch RB, Atmar RL, Franco LM, et al. Antibody correlates and predictors of immunity to naturally occurring influenza in humans and the importance of antibody to the neuraminidase. J Infect Dis 2013; 207:974–981.

28. Monto AS, Petrie JG, Cross RT, et al. Antibody to Influenza Virus Neuraminidase: An Independent Correlate of Protection. J Infect Dis 2015; 212:1191–1199.

29. Daulagala P, Mann BR, Leung K, et al. Imprinted Anti-Hemagglutinin and Anti-Neuraminidase Antibody Responses after Childhood Infections of A(H1N1) and A(H1N1)pdm09 Influenza Viruses. mBio 2023; 14:e0008423.

30. Zhang X, Ross TM. Anti-neuraminidase immunity in the combat against influenza. Expert Review of Vaccines 2024; 23:474–484.

31. Johansson BE, Kilbourne ED. Dissociation of influenza virus hemagglutinin and neuraminidase eliminates their intravirionic antigenic competition. J Virol 1993; 67:5721–5723.

32. Liang Z, Lin X, Sun L, et al. A(H2N2) and A(H3N2) influenza pandemics elicited durable cross-reactive and protective antibodies against avian N2 neuraminidases. Nat Commun 2024; 15:5593.

33. Chen Y, Wohlbold TJ, Zheng N-Y, et al. Influenza infection in humans induces broadly cross-reactive and protective neuraminidase-reactive antibodies. Cell 2018; 173:417–29.e10.

34. Easterbrook JD, Schwartzman LM, Gao J, et al. Protection against a lethal H5N1 influenza challenge by intranasal immunization with virus-like particles containing 2009 pandemic H1N1 neuraminidase in mice. Virology 2012; 432:39–44.

35. Rockman S, Brown LE, Barr IG, et al. Neuraminidase-Inhibiting Antibody Is a Correlate of Cross-Protection against Lethal H5N1 Influenza Virus in Ferrets Immunized with Seasonal Influenza Vaccine. J Virol 2013; 87:3053–3061.

36. Sun X, Belser JA, Li Z-N, et al. Effect of Prior Influenza A(H1N1)pdm09 Virus Infection on Pathogenesis and Transmission of Human Influenza A(H5N1) Clade 2.3.4.4b Virus in Ferret Model. Emerg Infect Dis 2025; 31:458–466.

37. Jiang L, Changsom D, Lerdsamran H. Cross-reactive antibodies against H7N9 and H5N1 avian influenza viruses in Thais population. Asian Pac J Allergy Immunol 2017; Available at: http://thailand.digitaljournals.org/index.php/APJAI/article/view/30689. Accessed 21 April 2025.

38. Daulagala P, Cheng SMS, Chin A, et al. Avian Influenza A(H5N1) Neuraminidase Inhibition Antibodies in Healthy Adults after Exposure to Influenza A(H1N1)pdm09. Emerg Infect Dis 2024; 30:168–171.

39. Pichyangkul S, Krasaesub S, Jongkaewwattana A, et al. Pre-Existing Cross-Reactive Antibodies to Avian Influenza H5N1 and 2009 Pandemic H1N1 in US Military Personnel. The American Society of Tropical Medicine and Hygiene 2014; 90:149–152.

40. Li Z-N, Liu F, Jung Y-J, et al. Population Immunity to Hemagglutinin Head, Stalk and Neuraminidase of Highly Pathogenic Avian Influenza 2.3.4.4b A(H5N1) viruses in the United States and the Impact of Seasonal Influenza on A(H5N1) Immunity. medRxiv preprint 2025; Available at: http://medrxiv.org/lookup/doi/10.1101/2025.03.30.25323419. Accessed 7 September 2025.

41. Ort JT, Leonard AS, Li SH, et al. Cross-reactive human antibody responses to H5N1 influenza virus neuraminidase are shaped by immune history. medRxiv preprint 2025; Available at: http://medrxiv.org/lookup/doi/10.1101/2025.09.02.25334929. Accessed 7 September 2025.

42. Garg S, Reinhart K, Couture A, et al. Highly Pathogenic Avian Influenza A(H5N1) Virus Infections in Humans. N Engl J Med 2024;

43. Mellis AM, Coyle J, Marshall KE, et al. Serologic Evidence of Recent Infection with Highly Pathogenic Avian Influenza A(H5) Virus Among Dairy Workers — Michigan and Colorado, June–August 2024. MMWR Morb Mortal Wkly Rep 2024; 73:1004–1009.

44. Werner AP, Schneider CG, Akin EH, et al. Low levels of H5N1 HA and NA antibodies in the human population are boosted by seasonal A/H1N1 infection but not by A/H3N2 infection or influenza vaccination. biorxiv preprint 2025; Available at: http://biorxiv.org/lookup/doi/10.1101/2025.07.13.664638. Accessed 8 September 2025.

45. Nuwarda RF, Alharbi AA, Kayser V. An Overview of Influenza Viruses and Vaccines. Vaccines 2021; 9:1032.

46. Bodewes R, de Mutsert G, van der Klis FRM, et al. Prevalence of antibodies against seasonal influenza A and B viruses in children in Netherlands. Clin Vaccine Immunol 2011; 18:469–476.

