## Supplementary Material for "Cross-reactive neuraminidase inhibition antibodies against H5N1 by consecutive influenza A imprinting cohorts of the past century: population-based serosurvey, British Columbia, Canada"

**Table S1.** Relatedness between hemagglutinin and neuraminidase proteins of H5N1 and representative human influenza A viruses since 1918

| Human influenza A subtype (HA, NA group), span | Period | Number of included HA / NA sequences[1] | H2N2 1957 pandemic strain[2] | | | H3N2 1968 pandemic, study virus[3] | | | H1N1 1977 pseudo-pandemic strain[4] | | | H1N1pdm09 2009 pandemic, study virus[5] | | | H5N1 2022 avian influenza, study virus[6,7,8] | | |
|---|---|---|---|---|---|---|---|---|---|---|---|---|---|---|---|---|---|
| | | | HA1 | HA2 | NA | HA1 | HA2 | NA | HA1 | HA2 | NA | HA1 | HA2 | NA | HA1 | HA2 | NA |
| H1N1 (1,1) 1918-1956 | 1918[9] | 1 | 60 | 84 | 45 | 37 | 58 | 45 | 82 | 94 | 87 | 83 | 92 | 89 | 56 | 83 | 95 |
| | 1919-1933 A-swine | 5 / 9 | 57-58 | 81-82 | 42-43 | 35-36 | 56 | 44 | 83-85 | 95-97 | 88-90 | 75-76 | 90-91 | 84-85 | 53-54 | 81-83 | 88-89 |
| | 1934-1946 A0 | 17 / 46 | 56-59 | 79-83 | 42-43 | 34-35 | 56-57 | 42-43 | 85-94 | 95-98 | 89-93 | 73-78 | 90-92 | 84-86 | 52-56 | 80-83 | 88-90 |
| | 1947-1956 A-prime | 23 / 22 | 56-58 | 82-84 | 42-44 | 33-35 | 56 | 43-44 | 91-99 | 96-99 | 91-97 | 72-77 | 90-92 | 84-86 | 52-54 | 81-83 | 86-89 |
| H2N2 (1,2) 1957-1967 | 1957[2] | 1 | 100 | 100 | 100 | 38 | 54 | 95 | 56 | 81 | 45 | 55 | 81 | 46 | 67 | 88 | 46 |
| | 1958-1967 | 42 / 18 | 92-100 | 96-100 | 95-99 | 37-38 | 52-55 | 94-98 | 53-57 | 79-83 | 42-43 | 53-55 | 80-83 | 44-45 | 65-68 | 85-88 | 43-44 |
| H3N2 (2,2) 1968-ongoing | 1968[3] | 1 | 38 | 54 | 95 | 100 | 100 | 100 | 35 | 55 | 44 | 38 | 58 | 46 | 36 | 54 | 46 |
| | 1969-2021 | 67,083 / 56,585 | 30-36 | 46-55 | 81-100 | 74-100 | 83-100 | 80-95 | 29-34 | 46-55 | 43-46 | 32-37 | 49-58 | 44-46 | 30-35 | 47-55 | 43-46 |
| | 2022[10] | 1 | 35 | 52 | 83 | 80 | 94 | 84 | 33 | 52 | 45 | 37 | 55 | 46 | 35 | 52 | 47 |
| H1N1 (1,1) 1977-2008 | 1977[4] | 1 | 56 | 81 | 45 | 35 | 55 | 44 | 100 | 100 | 100 | 72 | 91 | 83 | 52 | 80 | 86 |
| | 1978-2008 | 1,791 / 2,538 | 55-57 | 81-84 | 41-44 | 33-35 | 54-57 | 42-44 | 94-100 | 97-98 | 87-98 | 71-74 | 88-92 | 82-86 | 51-54 | 80-84 | 86-89 |
| H1N1pdm09 (1,1) 2009-ongoing | 2009[5] | 1 | 55 | 81 | 46 | 38 | 58 | 46 | 72 | 91 | 83 | 100 | 100 | 100 | 53 | 83 | 92 |
| | 2010-2021 | 44,275 / 41,380 | 52-56 | 76-82 | 43-46 | 33-38 | 55-59 | 43-45 | 67-75 | 85-92 | 80-88 | 90-100 | 93-100 | 90-100 | 50-54 | 77-84 | 86-93 |
| | 2022[11] | 1 | 56 | 82 | 46 | 38 | 58 | 45 | 71 | 89 | 83 | 91 | 97 | 95 | 53 | 82 | 88 |

Displayed are the range of percent amino acid pairwise identities within hemagglutinin (HA), including HA head (HA1) and HA stalk (HA2), and neuraminidase (NA) head domains of representative strains of influenza A subtypes. Pandemic years are red shaded. After removing sequences with duplicate names, incomplete coverage, swine origin or products of molecular cloning, percent pairwise identities in matched HA1, extracellular HA2 and NA head domains were derived as the number of identical amino acid residues between paired sequences divided by the total number of aligned amino acid positions. See reference citation for other identity comparisons,[12] including other H5N1 viruses. See **Table S2** for GISAID identifiers.[13]

---

[1] All protein sequences were downloaded from the Global Initiative on Sharing All Influenza Data (GISAID), on June 11, 2024 spanning sample collection dates from January 1, 1918 to December 31, 2022
[2] H2N2 pandemic strain, represented by A/Rotterdam/1957
[3] H3N2 pandemic strain represented by study virus: A/Hong Kong/1/1968.
[4] H1N1 pseudo-pandemic seasonal strain represented by: A/USSR/90/1977
[5] H1N1pdm09 pandemic strain representative of study virus: A/Mexico/InDRE4487/2009
[6] H5N1 belongs to HA group 1 and NA group 1
[7] H5N1 clade 2.3.4.4b avian influenza study virus: A/RT-Hawk/ON/FAV-0473-4/2022.
[8] H5N1 study virus shares >95% pairwise N1 head amino acid identity with representative clade 2.3.4.4b H5N1 viruses from recent human cases including bovine- (e.g. Texas/37/2024) and avian-origin (e.g., A/Ohio/06-1/2025, British Columbia/PHL-2032/2024).
[9] H1N1 pandemic strain represented by: A/Washington/NIAID-001-1918
[10] H3N2 contemporary seasonal strain represented by study virus: A/Massachusetts/18/2022 (recommended 2024-25 cell-based vaccine strain)
[11] H1N1pdm09 contemporary seasonal strain represented by study virus: A/Wisconsin/67/2022 (recommended 2023-24 and 2024-25 cell-based vaccine strain)
[12] Skowronski DM, Kaweski SE, Separovic L, *et al.* Neuraminidase imprinting and the age-related risk of zoonotic influenza. 2025; published online July 5. *medRxiv preprint*. DOI:10.1101/2025.07.03.25330844.
[13] Shu Y, McCauley J. GISAID: Global initiative on sharing all influenza data - from vision to reality. *Euro Surveill* 2017; **22**: 30494.

**Table S2.** Hemagglutinin and neuraminidase protein sequences, study and reference viruses, from GISAID

| HA Segment ID | NA Segment ID | Country | Collection Date | Isolate ID | Isolate Name | Originating Lab | Submitting Lab | Authors |
|---|---|---|---|---|---|---|---|---|
| EPI2971423 | EPI2971421 | United States | 1918-Sept-29 | EPI_ISL_18846999 | A/Washington/NIAID-001/1918 | National Institute of Allergy and Infectious Diseases | National Institute of Allergy and Infectious Diseases | Xiao, Y |
| EPI297875 | EPI297877 | Netherlands | 1957 | EPI_ISL_84911 | A/Rotterdam/1957 | | | Fouchier, R |
| EPI542494 | EPI542508 | Netherlands | 1968-Feb-29 | EPI_ISL_166563 | A/Netherlands/B1/1968 | | | Linster,M.; Lexmond,P.; Bestebroer,T.; Osterhaus,A.; Fouchier,R.; Herfst,S. |
| EPI901058 | EPI901060 | China | 1968 | EPI_ISL_245769 | A/Hong Kong/001/1968 | | | Lohrmann,F.; Dijkman,R.; Stertz,S.; Thiel,V.; Haller,O.; Staeheli,P.; Kochs,G. |
| EPI2096756 | EPI2096755 | United States | 2022-Jun-04 | EPI_ISL_13897304 | A/Massachusetts/18/2022 | Massachusetts Department of Public Health | Centers for Disease Control and Prevention | |
| EPI241092 | EPI241094 | Russian Federation | 1977 | EPI_ISL_69311 | A/USSR/90/1977 | | | Cummings,I.W.; Salser,W.A. |
| EPI178281 | EPI178287 | Mexico | 2009-Apr-14 | EPI_ISL_29924 | A/Mexico/InDRE4487/2009 | | National Microbiology Laboratory, Public Health Agency of Canada* | Bastien,N., Graham,M., Tyler,S., Van Domselaar,G., Drebot,M., Plummer,F., Aranda,C.A., Zavala,E.P., Eshaghi,A., Gubbay,J., Guyard,C., Guthrie,J., Duncan,C., Elgngihy,N., Tijet,N., Farrell,D., Drews,S., Hatchette,T., Davidson,R., Sarwal,S., Watson-Creed,G., Preiksaitis,J., Pabbaraju,K., Wong,S. and Li,Y.* |
| EPI2224978 | EPI2224977 | United States | 2022-Oct-22 | EPI_ISL_15928563 | A/Wisconsin/67/2022 | Wisconsin State Laboratory of Hygiene | Centers for Disease Control and Prevention | |
| EPI2500041 | EPI2500043 | Canada | 2022-Apr-26 | EPI_ISL_17394087 | A/RT-Hawk/ON/FAV-0473-4/2022 | Ontario Veterinary College, University of Guelph | Canadian Food Inspection Agency | Alkie, T; Ojkic, D; Stevens, B; Xu, W; Hisanaga, T; Koziuk, T; Fisher, M; Lung, O; Berhane, Y |
| EPI3650014 | EPI3650016 | Canada | 2024-Nov-09 | EPI_ISL_19548836 | A/British_Columbia/PHL-2032/2024 | British Columbia Centre for Disease Control (BCCDC); Public Health Agency of Canada (PHAC) | B.C. Centre for Disease Control | Shannon Russell, Natalie Prystajecky, Linda Hoang, Agatha Jassem, James Zlosnik, John Tyson, Frankie Tsang, Jessica Caleta, John Palmer, Dan Fornika, Kevin Yang, Kevin Kuchinski, Tracy Lee, Rob Azana, Janet Fung, Michael Chan, Branco Cheung, Nathalie Bastien, Ruimin Gao, Cody Buchanan, Jasmine Frost, Taeyo Chestley, Charlene Ranadheera |
| EPI3171488 | EPI3171486 | United States | 2024-Mar-28 | EPI_ISL_19027114 | A/Texas/37/2024 | Texas Department of State health Services – Laboratory Services | Centers for Disease Control and Prevention | Presley, Steven M; Webb, Cynthia Reinoso; Malaeb, Sierra; Cofi, Malory; Davis, Todd; Kondor, Becky; Steel, John; Kirby, Marie; Sheffield, Sydney; Liddell, Jimma; Frederick, Julia; Di, Han; Pusch, Elizabeth; Lacek, Kristine; Barnes, John |
| EPI4118067 | EPI4118066 | United States | 2025-Feb-12 | EPI_ISL_19785793 | A/Ohio/06-1/2025 | Ohio Department of Health Laboratories | Centers for Disease Control and Prevention | Dugan, Vivien; Davis, Todd; Kondor, Becky; Kirby, Marie; De La Cruz, Juan; Di, Han; Wilson, Malania; Sheffield, Sydney; Liddell, Jimma; Frederick, Julia; Lacek, Kristine; Keong, Lisa; Shu, Bo; DaSilva, Juliana; Ford, Jared; Johnson, Ashley |

GISAID = Global Initiative on Sharing All Influenza Data[1]; HA = hemagglutinin; NA = neuraminidase
*Data from matching GenBank entry, reference IDs: FJ998208.1 (HA) and FJ998214.1 (NA)

[1] Shu Y, McCauley J. GISAID: Global initiative on sharing all influenza data - from vision to reality. *Euro Surveill* 2017; **22**: 30494.

**Table S3**. GMTs and percent meeting NAI threshold titres against H5N1 and 2009-H1N1pdm09, by age group and sex

| Age group[1] (years) | Anti-N1 against H5N1[2] | | | | | | Anti-N1 against 2009-H1N1pdm09[1] | | | | | |
|---|---|---|---|---|---|---|---|---|---|---|---|---|
| | N | GMTs (95%CI)[3] | ≥10 n (%) (95%CI) | ≥40 n (%) (95%CI)[4] | ≥80 n (%) (95%CI)[4] | ≥160 n (%) (95%CI)[4] | N | GMTs (95%CI)[3] | ≥10 n (%) (95%CI)[4] | ≥40 n (%) (95%CI)[5] | ≥80 n (%) (95%CI)[4] | ≥160 n (%) (95%CI)[4] |
| 1-4 | 50 | 6.0 (5.2, 6.9) | 7 (14.0) (5.8, 26.7) | 0 (0) (0, 7.1) | 0 (0) (0, 7.1) | 0 (0) (0, 7.1) | 12 | 15.5 (5.2, 46.1) | 4 (33.3) (9.9, 65.1) | 4 (33.3) (9.9, 65.1) | 3 (25.0) (5.5, 57.2) | 2 (16.7) (2.1, 48.4) |
| 5-9 | 50 | 7.8 (6.0, 10.0) | 14 (28.0) (16.2, 42.5) | 3 (6.0) (1.3, 16.6) | 1 (2.0) (0.1, 10.7) | 1 (2.0) (0.1, 10.7) | 13 | 26.9 (7.8, 92.1) | 6 (46.2) (19.2, 74.9) | 6 (46.2) (19.2, 74.9) | 5 (38.5) (13.9, 68.4) | 2 (15.4) (1.9, 45.4) |
| 10-19 | 96 | 43.7 (31.4, 60.8) | 77 (80.2) (70.8, 87.6) | 47 (49.0) (38.6, 59.4) | 33 (34.4) (25.0, 44.8) | 20 (20.8) (13.2, 30.3) | 50 | 289.0 (166.9, 500.4) | 44 (88.0) (75.7, 95.5) | 43 (86.0) (73.3, 94.2) | 38 (76.0) (61.8, 86.9) | 36 (72.0) (57.5, 83.8) |
| 20-29 | 74 | 107.3 (77.8, 147.9) | 70 (94.6) (86.7, 98.5) | 57 (77.0) (65.8, 86.0) | 43 (58.1) (46.1, 69.5) | 29 (39.2) (28.0, 51.2) | 38 | 328.7 (190.4, 567.4) | 35 (92.1) (78.6, 98.3) | 34 (89.5) (75.2, 97.1) | 34 (89.5) (75.2, 97.1) | 30 (78.9) (62.7, 90.4) |
| 30-39 | 50 | 64.1 (43.2, 95.0) | 42 (84.0) (70.9, 92.8) | 35 (70.0) (55.4, 82.1) | 27 (54.0) (39.3, 68.2) | 13 (26.0) (14.6, 40.4) | 12 | 116.7 (47.5, 287.0) | 11 (91.7) (61.5, 99.8) | 10 (83.3) (51.6, 97.9) | 7 (58.3) (27.7, 84.8) | 7 (58.3) (27.7, 84.8) |
| 40-49 | 50 | 45.7 (31.6, 66.2) | 43 (86.0) (73.3, 94.2) | 28 (56.0) (41.3, 70.0) | 16 (32.0) (19.5, 46.7) | 8 (16.0) (7.2, 29.1) | 25 | 90.9 (46.3, 178.4) | 22 (88.0) (68.8, 97.5) | 18 (72.0) (50.8, 87.9) | 15 (60.0) (38.7, 78.9) | 9 (36.0) (18.0, 57.5) |
| 50-59 | 51 | 25.6 (17.6, 37.3) | 36 (70.6) (56.2, 82.5) | 20 (39.2) (25.8, 53.9) | 12 (23.5) (12.8, 37.5) | 3 (5.9) (1.2, 16.2) | 26 | 92.6 (44.9, 190.9) | 22 (84.6) (65.1, 95.6) | 19 (73.1) (52.2, 88.4) | 16 (61.5) (40.6, 79.8) | 9 (34.6) (17.2, 55.7) |
| 60-69 | 52 | 11.4 (8.4, 15.6 | 24 (46.2) (32.2, 60.5) | 8 (15.4) (6.9, 28.1) | 4 (7.7) (2.1, 1835) | 2 (3.8) (0.5, 13.2) | 26 | 25.5 (13.3, 48.7) | 16 (61.5) (40.6, 79.8) | 11 (42.3) (23.4, 63.1) | 7 (26.9) (11.6, 47.8) | 3 (11.5) (2.4, 30.2) |
| 70-79 | 52 | 42.4 (28.2, 63.7) | 43 (82.7) (69.7, 91.8) | 26 (50.0) (35.8, 64.2) | 18 (34.6) (22.0, 49.1) | 8 (15.4) (6.9, 28.1) | 27 | 274.8 (123.0, 613.7) | 25 (92.6) (75.7, 99.1) | 21 (77.8) (57.7, 91.4) | 20 (74.1) (53.7, 88.9) | 16 (59.3) (38.8, 77.6) |
| 80+ | 50 | 91.5 (62.0, 134.9) | 48 (96.0) (86.3, 99.5) | 36 (72.0) (57.5, 83.8) | 28 (56.0) (41.3, 70.0) | 14 (28.0) (16.2, 42.5) | 21 | 436.8 (236.1, 808.1) | 21 (100) (83.9, 100) | 20 (95.2) (76.2, 99.9) | 18 (85.7) (63.7, 97.0) | 18 (85.7) (63.7, 97.0) |
| **Overall** | 575 | 33.1 (29.0, 27.8) | 404 (70.3) (66.3, 74.0) | 260 (45.2) (41.1, 49.4) | 182 (31.7) (27.9, 35.6) | 98 (17.0) (14.1, 20.4) | 250 | 137.2 (106.7, 176.4) | 206 (82.4) (77.1, 86.9) | 186 (74.4) (68.5, 79.7) | 163 (65.2) (58.9, 71.1) | 132 (52.8) (46.4, 59.1) |
| **Age-standardized**[6] | 575 | n/a | (74.8) (71.4, 78.3) | (50.2) (46.1, 54.3) | (35.0) (30.9, 39.1) | (18.5) (15.1, 21.9) | 250 | n/a | (82.7) (77.9, 87.5) | (73.7) (67.8, 79.6) | (62.3) (55.6, 69.1) | (49.6) (42.8, 56.3) |
| **Sex** | | | | | | | | | | | | |
| Female | 287 | 36.7 (30.3, 44.4) | 208 (72.5) (66.9, 77.6) | 137 (47.7) (41.8, 53.7) | 96 (33.4) (28.0, 39.2) | 52 (18.1) (13.8, 23.1) | 125 | 109.1 (75.9, 156.9) | 100 (80.0) (71.9, 86.6) | 87 (69.6) (60.7, 77.5) | 77 (61.6) (52.5, 70.2) | 59 (47.2) (38.2, 56.3) |
| Male | 288 | 29.9 (25.0, 35.9) | 196 (68.1) (62.3, 73.4) | 123 (42.7) (36.9, 48.7) | 86 (29.9) (24.6, 35.5) | 46 (16.0) (11.9, 20.7) | 125 | 172.5 (121.8, 244.2) | 106 (84.8) (77.3, 90.6) | 99 (79.2) (71.0, 85.9) | 86 (68.8) (59.9, 76.8) | 73 (58.4) (49.2, 67.1) |

GMTs = geometric mean titres; NAI = Neuraminidase inhibition; n/a = not applicable

[1] With random selection of sera from among children <5 years, none <1 year was randomly chosen
[2] Test virus is a reassortment strain comprised of the H7 from A/Anhui/2013 (H7N9), the six internal proteins from A/PuertoRico/8/1934 (H1N1), and the neuraminidase from the specified study virus with details provided in the main manuscript, and footnotes, **Table S1**
[3] Significant variation in GMT by age group for both viruses, p<0.0001; no statistically significant variation in GMT by sex, p=0.13 for H5N1, p=0.07 for 2009-H1N1pdm09
[4] One-sided 97.5% confidence intervals provided for cells with a count of zero or 100%
[5] One-sided 97.5% confidence intervals provided for cells with a count of zero or 100%
[6] Directly age-standardized for the age distribution of the Lower Mainland area, British Columbia, Canada as per: Population projections. Vancouver: Government of British Columbia. Available: https://www2.gov.bc.ca/gov/content/data/statistics/people-population-community/population/population-projections (accessed 15 May 2025)

**Table S4**. GMTs and percent meeting NAI threshold titres against H5N1 and H1N1pdm09, by birth cohort

| Birth cohorts (years) | Sample size, by test virus | | Anti-N1 titres by test virus[1] | | | | | | | | | | | |
|---|---|---|---|---|---|---|---|---|---|---|---|---|---|---|
| | | | GMT (95% CI) | | | ≥40 n (%) (95%CI)[2] | | | ≥80 n (%) (95%CI)[2] | | | ≥160 n (%) (95%CI)[2] | | |
| | H5N1 | H1N1 pdm09[3,4] | H5N1[5] | 2009-H1N1 pdm09[4] | 2022-H1N1 pdm09[4] | H5N1 | 2009-H1N1 pdm09 | 2022-H1N1 pdm09 | H5N1 | 2009-H1N1 pdm09 | 2022-H1N1 pdm09 | H5N1 | 2009-H1N1 pdm09 | 2022-H1N1 pdm09 |
| 2020-2023 | 50 | 12 | 6.0 (5.2, 6.9) | 15.5 (5.2, 46.1) | 77.2 (13.7, 435.5) | 0 (0) (0, 7.1) | 4 (33.3) (9.9, 65.1) | 6 (50.0) (21.1, 78.9) | 0 (0) (0, 7.1) | 3 (25.0) (5.5, 57.2) | 6 (50.0) (21.1, 78.9) | 0 (0) (0, 7.1) | 2 (16.7) (2.1, 48.4) | 6 (50.0) (21.1, 78.9) |
| 2015-2019 | 50 | 13 | 7.8 (6.0, 10.0) | 26.9 (7.8, 92.1) | 165.8 (42.7, 643.2) | 3 (6.0) (1.3, 16.6) | 6 (46.2) (19.2, 74.9) | 9 (69.2) (38.6, 90.9) | 1 (2.0) (0.1, 10.7) | 5 (38.5) (13.9, 68.4) | 8 (61.5) (31.6, 86.1) | 1 (2.0) (0.1, 10.7) | 2 (15.4) (45.5, 92.0) | 8 (61.5) (31.6, 86.1) |
| 2015-2023 | 100 | 25 | 6.8 (5.9, 7.9) | 20.7 (9.6, 44.7) | 114.9 (41.6, 317.5) | 3 (3.0) (0.6, 8.5) | 10 (40.0) (21.1, 61.3) | 15 (60.0) (38.7, 78.9) | 1 (1.0) (0.0, 5.5) | 8 (32.0) (15.0, 53.5) | 14 (56.0) (34.9, 75.6) | 1 (1.0) (0.0, 5.5) | 4 (16.0) (4.5, 36.1) | 14 (56.0) (34.9, 75.6) |
| 2009-2014 | 50 | 25 | 35.1 (21.1, 58.3) | 211.5 (94.8, 472.1) | 243.0 (125.1, 471.9) | 20 (40.0) (26.4, 54.8) | 21 (84.0) (63.9, 95.5) | 22 (88.0) (68.8, 97.5) | 14 (28.0) (16.2, 42.5) | 17 (68.0) (46.5, 85.1) | 19 (76.0) (54.9, 90.6) | 10 (20.0) (10.0, 33.7) | 16 (64.0) (42.5, 82.0) | 15 (60.0) (38.7, 78.9) |
| 2004-2008 | 50 | 25 | 59.1 (38.6, 90.6) | 394.8 (179.7, 867.3) | 111.7 (58.0, 214.8) | 30 (60.0) 45.2, 73.6) | 22 (88.0) (68.8, 97.5) | 21 (84.0) (63.9, 95.5) | 21 (42.0) (28.2, 56.8) | 21 (84.0) (63.9, 95.5) | 19 (76.0) (54.9, 90.6) | 11 (22.0) (11.5, 36.0) | 20 (80.0) (59.3, 93.2) | 13 (52.0) (31.3, 72.2) |
| 1997-2003 | 50 | 25 | 100.8 (71.0, 143.2) | 427.9 (231.1, 792.4) | 123.8 (67.0, 228.7) | 39 (78.0) (64.0, 88.5) | 23 (92.0) (74.0, 99.0) | 18 (72.0) (50.6, 87.9) | 28 (56.0) (41.3, 70.0) | 23 (92.0) (74.0, 99.0) | 16 (64.0) (42.5, 82.0) | 20 (40.0) (26.4, 54.8) | 21 (84.0) (63.9, 95.5) | 10 (40.0) (21.1, 61.3) |
| 1985-1996 | 70 | 25 | 77.0 (54.2, 109.2) | 153.6 (76.9, 306.7) | 54.5 (31.7, 93.8) | 50 (71.4) (59.4, 81.6) | 21 (84.0) (63.9, 95.5) | 16 (64.0) (42.5, 82.0) | 40 (57.1) (44.8, 68.9) | 18 (72.0) (50.6, 87.9) | 9 (36.0) (18.0, 57.5) | 21 (30.0) (19.6, 42.1) | 16 (64.0) (42.5, 82.0) | 6 (24.0) (9.4, 45.1) |
| 1977-1984 | 50 | 25 | 45.7 (31.6, 66.2) | 90.9 (46.3, 178.4) | 44.4 (25.9, 76.2) | 28 (56.0) (41.3, 70.0) | 18 (72.0) (50.6, 87.9) | 11 (44.0) (24.4, 65.1) | 16 (32.0) (19.5, 46.7) | 15 (60.0) (38.7, 78.9) | 8 (32.0) (15.0, 53.5) | 8 (16.0) (7.2, 29.1) | 9 (36.0) (18.0, 57.5) | 4 (16.0) (4.5, 36.1) |
| 1968-1976 | 50 | 25 | 25.7 (17.6, 37.7) | 95.1 (44.8, 201.8) | 31.5 (15.8, 62.5) | 20 (40.0) (26.4, 54.8) | 18 (72.0) (50.6, 87.9) | 9 (36.0) (18.0, 57.5) | 12 (24.0) (13.1, 38.2) | 16 (64.0) (42.5, 82.0) | 7 (28.0) (12.1, 49.4) | 3 (6.0) (1.3, 16.6) | 9 (36.0) (18.0, 57.5) | 6 (24.0) (9.4, 45.1) |
| 1957-1967 | 50 | 25 | 10.7 (8.0, 14.2) | 25.1 (13.4, 46.8) | 17.1 (9.7, 30.2) | 7 (14.0) (5.8, 26.7) | 11 (44.0) (24.4, 65.1) | 8 (32.0) (15.0, 53.5) | 7 (6.0) (1.3, 16.6) | 6 (24.0) (9.4, 45.1) | 5 (20.0) (6.8, 40.7) | 1 (2.0) (0.1, 10.7) | 2 (8.0) (1.0, 26.0) | 1 (4.0) (0.1, 20.4) |
| 1947-1956 | 50 | 25 | 44.7 (29.3, 68.4) | 248.2 (100.5, 612.9) | 67.7 (34.3, 133.7) | 26 (52.0) (37.4, 66.3) | 19 (76.0) (54.9, 90.6) | 13 (52.0) (31.3, 72.2) | 18 (36.0) (22.9, 50.8) | 18 (72.0) (50.6, 87.9) | 9 (36.0) (18.0, 57.5) | 8 (16.0) (7.2, 29.1) | 15 (60.0) (38.7, 78.9) | 8 (32.0) (15.0, 53.5) |
| Pre-1947 | 55 | 25 | 81.0 (55.4, 118.6) | 387.3 (218.4, 686.6) | 74.9 (41.5, 135.1) | 37 (67.3) (53.3, 79.3) | 23 (92.0) (74.0, 99.0) | 17 (68.0) (46.5, 85.1) | 29 (52.7) (38.8, 66.4) | 21 (84.0) (63.9, 95.5) | 11 (44.0) (24.4, 65.1) | 15 (27.3) (16.1, 41.0) | 20 (80.0) (59.3, 93.2) | 9 (36.0) (18.0, 57.5) |

GMT = geometric mean titre; NAI = Neuraminidase inhibition

[1] All test viruses are reassortment strains comprised of the H7 from A/Anhui/2013 (H7N9), the six internal proteins from A/PuertoRico/8/1934 (H1N1), and the neuraminidase from study viruses with specifications provided in the main manuscript, and footnotes, **Table S1**
[2] One-sided 97.5% confidence intervals provided for cells with a count of zero
[3] Samples include 21 tested only for anti-N1 to H1N1pdm09 strains, including six each from the pre-1947 and 1947-56, seven from the 1997-2003 and two from the 2004-08 birth cohorts
[4] Sample sizes apply to both 2009-H1N1pdm09 and 2022-H1N1pdm09
[5] Significant variation in GMT by birth cohort, p<0.0001 with or without stratification among 2015-2023 birth cohort

**Table S5**. GMTs and percent meeting NAI threshold titres against H3N2, by birth cohort

| Selected birth cohorts (years) | Sample size | Anti-N2 by test virus[1] | | | | | | | |
|---|---|---|---|---|---|---|---|---|---|
| | | GMT (95% CI) | | ≥40 n (%) (95%CI)[2] | | ≥80 n (%) (95%CI)[2] | | ≥160 n (%) (95%CI)[2] | |
| | | 1968-H3N2[3] | 2022-H3N2[4] | 1968-H3N2 | 2022-H3N2 | 1968-H3N2 | 2022-H3N2 | 1968-H3N2 | 2022-H3N2 |
| 1997-2003 | 50 | 31.46 (21.6, 45.9) | 15.3 (11.6, 20.0) | 24 (48.0) (33.7, 62.6) | 9 (18.0) (8.6, 31.4) | 13 (26.0) (14.6, 40.4) | 1 (2.0) (0.05, 10.7) | 5 (10.0) (3.3, 21.8) | 1 (2.0) (0.05, 10.7 |
| 1968-1976[5] | 50 | 89.0 (66.4, 119.3) | 8.9 (7.1, 11.3) | 43 (86.0) (73.3, 94.2) | 4 (8.0) (2.2, 19,2) | 27 (54.0) (39.3, 68.2) | 1 (2.0) (0.05, 10.7) | 15 (30.0) (17.9, 44.6) | 0 (0.0) (0, 7.1) |
| 1957-1967 | 50 | 642.7 (509.1, 811.4) | 9.9 (7.1, 13.8) | 50 (100) (92.9, 100) | 6 (12.0) (4.53, 24.3) | 50 (100) (92.9, 100) | 3 (6.0) (1.3, 16.6) | 50 (100) (92.9, 100) | 2 (4.0) (0.49, 13.7) |
| Pre-1947 | 55 | 279.4 (228.3, 342.0) | 12.2 (9.1, 16.4) | 55 (100) (93.51, 100) | 9 (16.4) (7.8, 28.8) | 53 (96.4) (87.47, 99.6) | 5 (9.1) (3.0, 20.0) | 43 (78.2) (65.0, 88.2) | 1 (1.8) (0.05, 9.7) |

**Table S6**. GMTs and percent meeting NAI thresholds against H1N1pdm09, restricted to samples also tested for H3N2, by birth cohort

| Selected birth cohorts (years) | Sample size | Anti-N1 by test virus[1] | | | | | | | |
|---|---|---|---|---|---|---|---|---|---|
| | | GMT (95% CI) | | ≥40 n (%) (95%CI) | | ≥80 n (%) (95%CI) | | ≥160 n (%) (95%CI) | |
| | | 2009-H1N1pdm09 | 2022-H1N1pdm09 | 2009-H1N1pdm09 | 2022-H1N1pdm09 | 2009-H1N1pdm09 | 2022-H1N1pdm09 | 2009-H1N1pdm09 | 2022-H1N1pdm09 |
| 1997-2003 | 18 | 375.6 (169.8, 830.6) | 109.3 (50.0, 238.8) | 16 (88.9) (65.3, 98.6) | 12 (66.7) (41.0, 86.7) | 16 (88.9) (65.3, 98.6) | 11 (61.1) (35.8, 82.7) | 15 (83.3) (58.6, 96.4) | 7 (38.9) (17.3, 64.3) |
| 1968-1976[3] | 25[6] | 95.1 (44.8, 201.8) | 31.5 (15.8, 62.5) | 18 (72.0) (50.6, 87.9) | 9 (36.0) (18.0, 57.5) | 16 (64.0) (42.5, 82.0) | 7 (28.0) (12.1, 49.4) | 9 (36.0) (18.0, 57.5) | 6 (24.0) (9.4, 45.1) |
| 1957-1967 | 25[4] | 25.1 (13.4, 46.8) | 17.1 (9.7, 30.2) | 11 (44.0) (24.4, 65.1) | 8 (32.0) (15.0, 53.5) | 6 (24.0) (9.4, 45.1) | 5 (20.0) (6.8, 40.7) | 2 (8.0) (1.0, 26.0) | 1 (4.0) (0.1, 20.4) |
| Pre-1947 | 19 | 381.0 (183.4, 791.4) | 77.9 (35.8, 169.5) | 17 (89.47) (66.9, 98.7) | 12 (63.2) (38.4, 83.7) | 16 (84.2) (60.4, 96.6) | 10 (52.6) (28.9, 75.6) | 15 (78.9) (54.4, 94.0) | 8 (42.1) (20.3, 66.50) |

GMT = geometric mean titre; NAI = Neuraminidase inhibition

[1] All test viruses are reassortment strains comprised of the H7 from A/Anhui/2013 (H7N9), the six internal proteins from A/PuertoRico/8/1934 (H1N1), and the neuraminidase from study viruses by subtype, with specifications provided in main manuscript and footnotes, **Table S1**
[2] One-sided 97.5% confidence intervals provided for cells with 0% or 100%
[3] Significant variation in GMT by birth cohort, p<0.0001
[4] No statistically significant variation in GMT by birth cohort, p=0.044
[5] Birth year (age span) of tested samples 1968-1974 (50-56 years)
[6] All samples originally tested against H1N1pdm09 viruses were included in H3N2 testing; values carried over from **Table S4**

**Figure S1.** Scatterplot of NAI titres, H5N1 and 2022-H1N1pdm09

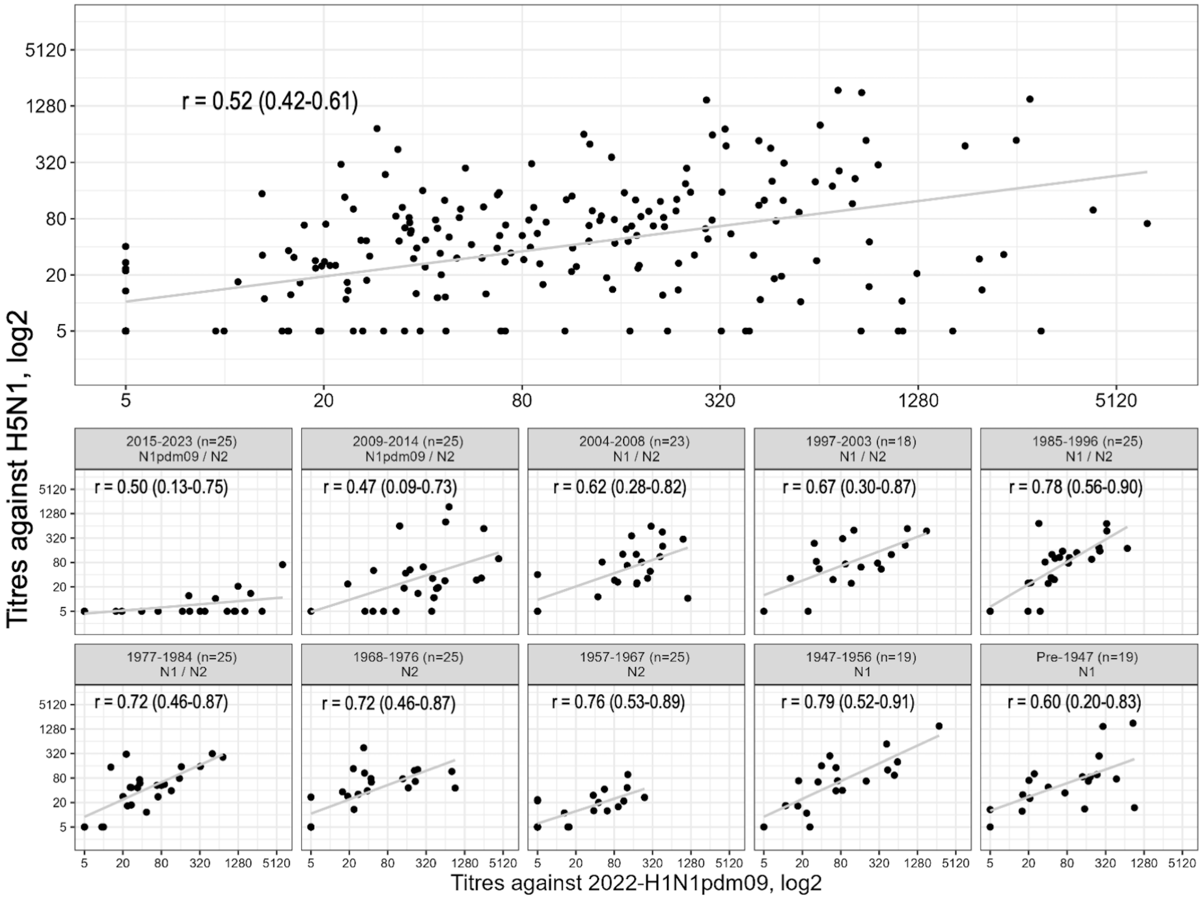

Scatterplots display the relationship between individual H5N1 and 2022-H1N1pdm09 neuraminidase inhibition (NAI) antibody titres, overall and by birth cohort, with accompanying Pearson correlation coefficients (r). Both test viruses are reassortment strains comprised of the H7 from A/Anhui/2013 (H7N9), the six internal proteins from A/PuertoRico/8/1934 (H1N1), and the neuraminidase from study viruses with specifications provided in footnotes, **Table S1**. Within panel headers by birth cohort are displayed the range of cohort birth years (sample size) and circulating neuraminidase subtype. Only individuals tested for both viruses are included (n=229).

**Figure S2.** Scatterplot of NAI titres, 2009-H1N1pdm09 and 1968-H3N2

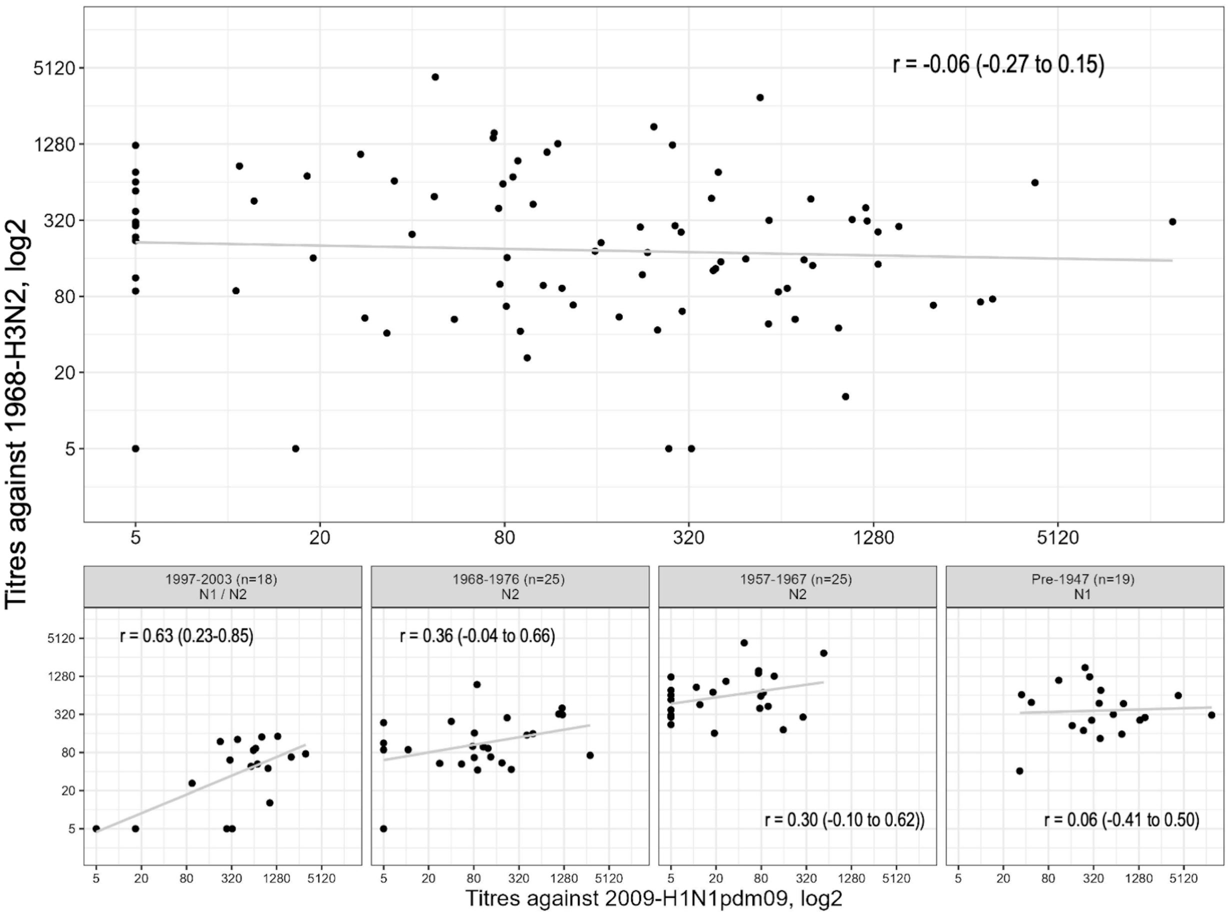

Scatterplots display the relationship between individual 2009-H1N1pdm09 and 1968-H3N2 neuraminidase inhibition (NAI) antibody titres overall and by birth cohort, with accompanying Pearson correlation coefficients (r). Both test viruses are reassortment strains comprised of the H7 from A/Anhui/2013 (H7N9), the six internal proteins from A/PuertoRico/8/1934 (H1N1), and the neuraminidase from study viruses with specifications provided in footnotes, **Table S1**. Within panel headers by birth cohort are displayed the range of cohort birth years (sample size) and circulating neuraminidase subtype. Only individuals tested for both viruses are included (n=87).

**Figure S3.** Scatterplot of NAI titres, H5N1 and 1968-H3N2

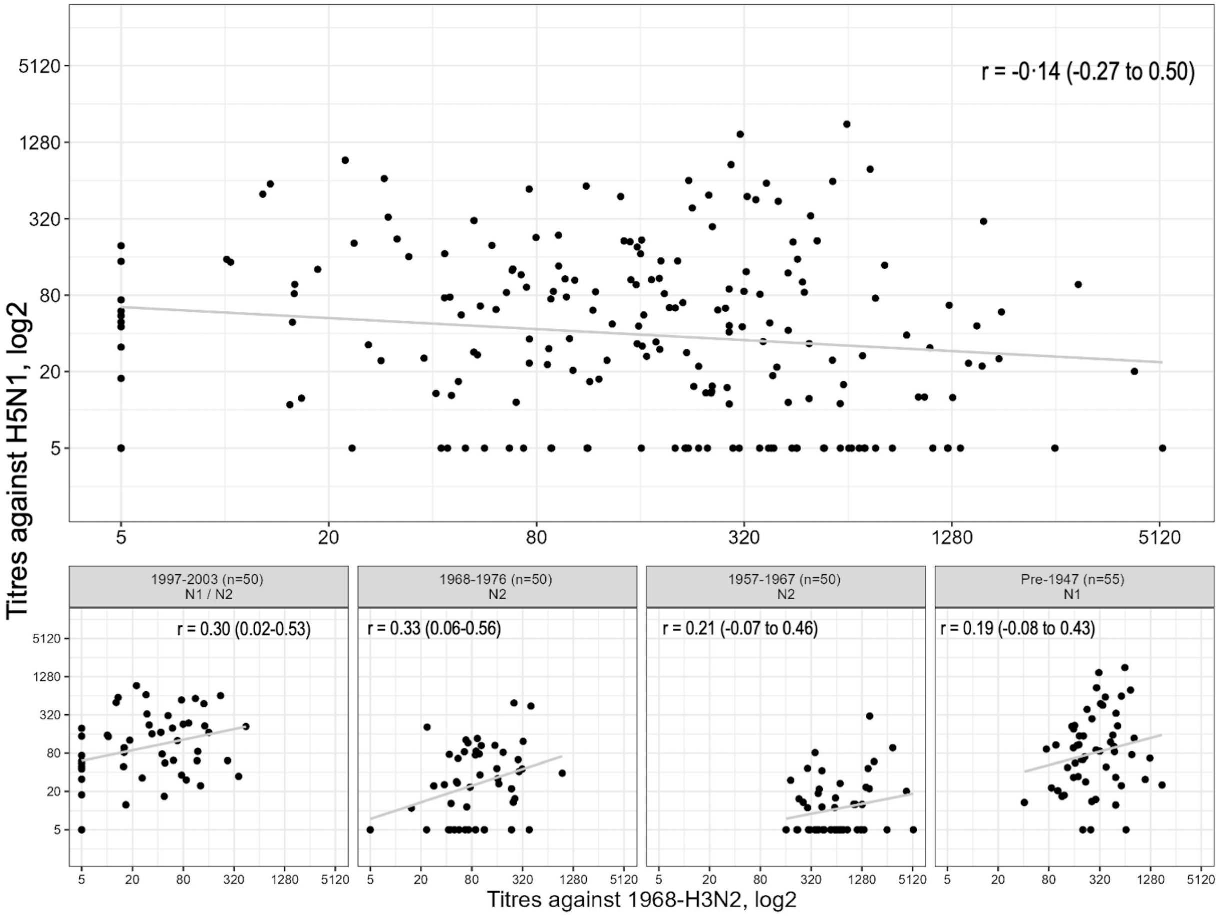

Scatterplots display the relationship between individual H5N1 and 1968-H3N2 neuraminidase inhibition (NAI) antibody titres overall and by birth cohort, with accompanying Pearson correlation coefficients (r). Both test viruses are reassortment strains comprised of the H7 from A/Anhui/2013 (H7N9), the six internal proteins from A/PuertoRico/8/1934 (H1N1), and the neuraminidase from study viruses with specifications provided in footnotes, **Table S1**. Within panel headers by birth cohort are displayed the range of cohort birth years (sample size) and circulating neuraminidase subtype. Only individuals tested for both viruses are included (n=205).
